# Experimental investigation of muscle-tendon unit geometry and kinematics in lower-limb muscles during gait: Current Applications and Future Directions – A Scoping Review

**DOI:** 10.1101/2025.09.16.25335871

**Authors:** Thomas Lecharte, Fabien Leboeuf, Raphaël Gross, Antoine Nordez, Guillaume Le Sant, Cloé Dussault-Picard

## Abstract

**Purpose:** Musculoskeletal (MSK) modeling and ultrasound imaging (USI) are complementary techniques that, when combined with three-dimensional gait analysis (3DGA), provide insights into muscle and/or muscle–tendon unit (MTU) characteristics during gait. Despite their potential, a synthesis of their current use during 3DGA in populations with neuromotor impairments has not been conducted. This scoping review aimed to examine how MSK modeling and USI are used alongside 3DGA to assess muscle and MTU characteristics in populations with neuromotor impairments and evaluate the potential clinical implications of these approaches on clinical assessment and decision-making.

**Results:** Three databases were searched up to February 2025, yielding 50 studies (42 studies used MSK modeling, 5 used USI, and 3 employed both), including 4 pathological populations (cerebral palsy, stroke, hereditary spastic paraparesis, and idiopathic toe walking), and analyzing 11 lower-limb muscles. Both MSK modeling and USI have enabled the assessment of muscle or MTU length during gait, and detection of abnormal MTU. Only MSK modeling was used to assess MTU lengthening velocity, interventions effects, and predictors of surgical outcomes. MSK modeling appears limited by modeling assumptions and lack of real-time data, whereas USI faces constraints related to data acquisition complexity and processing challenges.

**Conclusions:** This review enhances understanding of neuromuscular impairments and current uses of MSK modeling and USI in clinical populations. It highlights their complementary potential with 3DGA to support personalized clinical decision-making. Future work should include broader neuromotor conditions and explore automated data analysis (e.g., deep learning for USI) to improve clinical applicability.

## 1. Introduction

Three-dimensional gait analysis (3DGA), which assesses kinematics, kinematics, and muscle electromyography during gait, is widely recognized as the gold-standard clinical tool for evaluating gait [1]. 3DGA allows identification of gait deviations resulting from neuromusculoskeletal impairments [2], and contributes to therapeutic planning [3]. While 3DGA is highly valuable for detecting gait abnormalities, it offers limited insights on musculoskeletal parameters such as muscle and/or muscle-tendon unit (MTU) geometry (e.g., fascicle and MTU length, respectively), and muscle and/or MTU kinematics (e.g., fascicle and MTU lengthening velocity, respectively). Muscle specific parameters are critical for clinical decision-making such as muscle strengthening or selective lengthening of the tendon or muscle [4]. For instance, 3DGA often associates initial contact with a flexed knee to hamstrings shortening, frequently resulting in a recommendation for surgical lengthening [5]. However, the clinical benefits of this procedure remain modest, with partial improvement of the gait pattern [6]. A more precise assessment of muscle length during 3DGA appeared as a promising option for improving the targeting and effectiveness of such interventions [7].

For that purpose, 3D musculoskeletal (MSK) modeling during 3DGA has emerged. By integrating sensor-based measurements (e.g., joint angles and/or ground reaction forces and/or muscle activity) and neuro-mechanical algorithms, MSK modeling provides a comprehensive framework for assessing muscle and MTU geometry and kinematics [8,9]. Simplified 2D models often rely on geometric assumptions (e.g., regressive equations derived from cadavers anthropometric measurements and sagittal joint angles [10]), whereas advanced 3D frameworks incorporate detailed musculoskeletal geometries (e.g., bone and muscle structures reconstructed from imaging data) and patient-specific data (e.g., scaled musculoskeletal models based on subject-specific anthropometric data) for more accurate estimations (e.g., OpenSim [11])[12,13]. While MSK modeling offers insights into MTU geometry and kinematics, it does not capture the specific behavior of muscle fibers and tendons. Ultrasound imaging (USI) during gait has been used to address this gap. USI offers to visualize the muscle and/or its tendon during walking [14] by securing an ultrasound probe over the area of interest and synchronizing its data with a motion capture system [15]. It enables the quantification of muscle geometry such as pennation angle, muscle fiber length, fascicle length, and whole MTU length [16], and muscle-tendon junction (MTJ) displacement [17]. MSK modeling and USI are emerging non-invasive methods for assessing gait MTU geometry and kinematics, and the specific behavior of muscle fibers and tendons, respectively.

To the best of our knowledge, no comprehensive review has specifically focused on the application of these methods during 3DGA, and their clinical relevance in individuals with neuromotor impairments. Regarding MSK modeling during 3DGA, a recent systematic review has emphasized the importance of subject-specific scaled and personalized models (e.g., muscle insertion and pathway sites, joint centers) for more precise simulations [18]. Another systematic review also reported the reliability and challenges related to USI during gait [16], highlighting that both fascicle length and pennation angle can be measured reliably across and within session during dynamic tasks. However, the technical recommendations for more accurate estimations (e.g., scaling and personalizing MSK models [18], and securing the probe placement and adequate frame rate and filtering for USI [19]), have been predominantly derived from studies involving non-pathological, healthy individuals, potentially limiting their applicability in clinical populations. Yet, synthesizing clinical applications and potential interpretations in pathological populations is crucial to optimize the use of such technologies in clinical and research settings and to refine their indications. To address this gap, this scoping review aims (1) to examine how MSK modeling and USI are used in populations with neuromotor impairments during 3DGA to assess muscle and/or MTU geometry and/or kinematics, and (2) to explore their potential clinical implications in individuals with neuromotor impairments.

## 2. Methods

This scoping review follows the Systematic reviews and Meta-Analyses extension for Scoping Reviews (PRISMA-ScR) checklist [20] and was registered on the OSF platform (ID: 10.17605/OSF.IO/FXMKA). The question: “What are the current uses and clinical implications of MSK modeling and USI during gait in individuals with neuromotor impairments?” was addressed.

### 2.1. Eligibility criteria

The PCC (population, concept and context) model [21] was used to define the meaningful eligibility criteria of the included studies.

#### 2.1.1. Population

Studies were included if the population under investigation presented with a neuromotor impairment (e.g., stroke, incomplete spinal cord injury, cerebral palsy (CP)) and excluded if the population assessed did not present with neuromotor impairment or had another impairment.

#### 2.1.2. Concept

The current use and clinical implications of MSK modeling and USI were the main concept investigated. Studies were included if the outcomes were related to muscle and/or MTU geometry and/or kinematics (e.g., fascicle, muscle fiber, tendon, or MTU length and/or lengthening velocity and/or lengthening acceleration, pennation angle) and if it was conducted with USI and 3D MSK modeling. Thus, studies were excluded if the outcomes concerned other muscle and/or MTU parameters such as muscle lever arm, muscle activation, muscle force. Also, studies that made estimations through regression equations and mathematical model [10,22] were excluded, to retain studies relying on subject-specific or anatomically informed methods (i.e., MSK modeling or USI) and to avoid estimates by theoretical assumptions, which may not accurately reflect individual muscle behavior.

#### 2.1.3. Context

Studies were included if the outcomes of interest were assessed during forward leveled walking (e.g., laboratory or treadmill settings) and excluded if the task was not walking (e.g., running or stair climbing), and/or conducted on irregular or uneven surfaces.

#### 2.1.4. Type of studies

Studies were included if they were published in English as an original article. Therefore, studies written in other languages and reviews were excluded.

### 2.2. Data and literature sources

Studies were searched in PubMed, Scopus, and Clinical Trials in February 2025 based on 4 main concepts: “Experimental investigation”, “muscle-tendon unit geometry and/or kinematics”, “lower-limb muscles”, and “gait analysis” (see **supplementary file**).

### 2.3. Studies screening

The titles and abstracts were independently screened for eligibility based on the inclusion criteria by two authors (TL and CDP). Subsequently, a full-text screening of the selected studies was independently conducted by the same authors. In the case of disagreements, a third author (FL) achieved consensus. All the screening steps were conducted in Covidence (Veritas Health Innovation Ltd, Melbourne, Australia).

### 2.4. Methodological quality assessment

For each study, two authors (TL and CDP) conducted separate and blinded assessment of the methodological quality using the Joanna Briggs Institute critical appraisal checklists [21]. According to the study design, the appropriate checklist was selected. Based on the total number of criteria assessed according to the study type, each criterion was assigned a score of “yes” (1 pt) “unclear” (0.5 pt), “no” (0 pt), or not applicable (NA). A global score (%) of the study was then calculated:

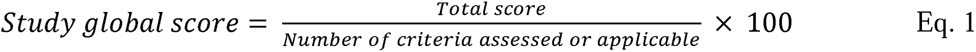

Before rating, the results obtained from 3 arbitrary studies were compared between both raters to ensure the standardization and reliability of the assessment. After rating all studies, both raters reached consensus on each criterion and the total score of each study.

### 2.5. Data charting process

Data including subject characteristics (age, sex, neuromotor impairment), study methods (walking speed, footwear condition, walking surface, number of gait trials or cycles, kinematic and musculoskeletal models used, normalization technique), outcomes (muscles assessed, muscle geometry and/or kinematics parameters, instant of the gait cycle assessed), and study aims and design were extracted by one author (TL or CDP), and validated by a second author (TL or CDP). Descriptive and numerical analyses were used to summarize the literature for each method (i.e., USI and MSK modeling). Then, according to the aim of this scoping review, the current use of MSK modeling and USI during gait (aim 1), along with their clinical contributions (aim 2) were discussed (**Figure 1)**.

**Figure 1.**
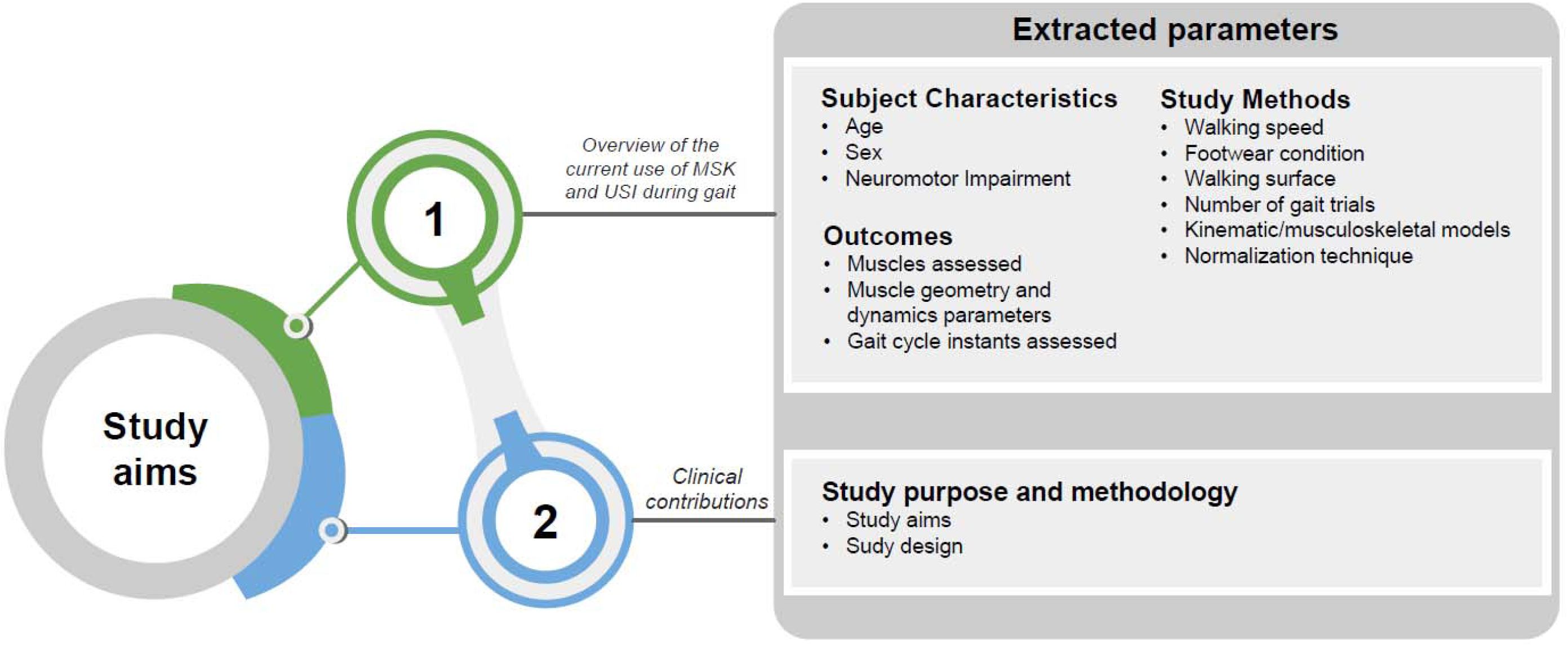
Representation of the data extracted to provide an overview of the current use of musculoskeletal modeling and ultrasound imaging during gait (aim 1), along with their clinical contributions (aim 2).

## 3. Results

No major deviation from the original registered protocol is to be reported. The search strategy identified 984 studies. The **Figure 2** illustrated the selection process using a PRISMA flowchart [23]. A total of 50 studies were eligible for this review [4,24–72]. Among them, 42 focused exclusively on MSK modeling [24–33,36–60,66–72] (n=2425 participants, 721 males/484 females/1220 not specified (n/s); mean age±SD = 10.9±4.1 years), 5 focused exclusively on USI [4,61–64] (n=109 participants, 62 males/47 females; mean age±SD = 18.1±5.5 years), and 3 focused on both MSK modelling and USI (n=87 participants, 27 males/26 females/4 n/s; mean age±SD = 11.0±3.4 years)[34,35,65].

**Figure 2.**
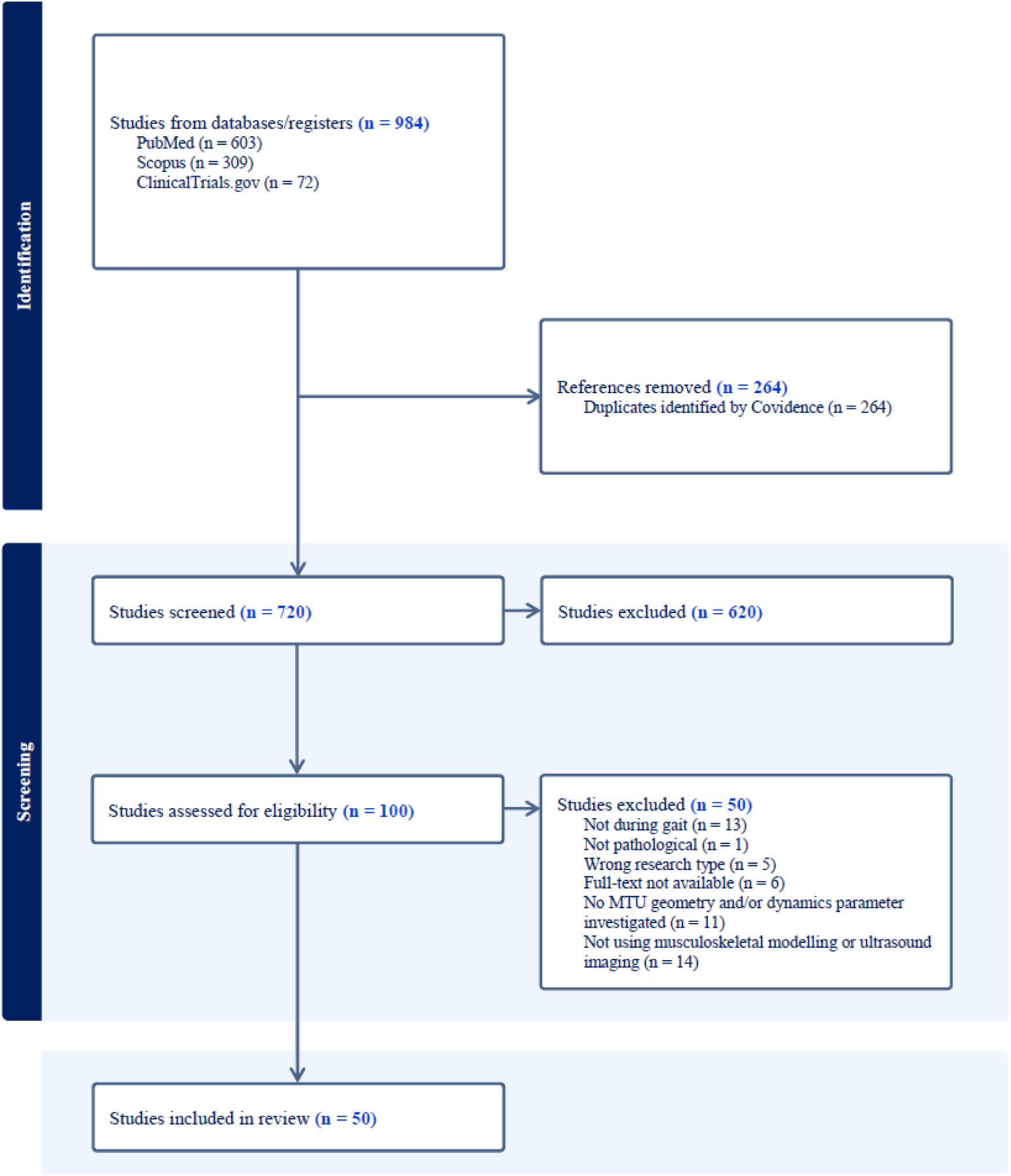
Prisma flowchart produced by Covidence software (Veritas Health Innovation Ltd, Melbourne, Australia) representing the study selection.

### 3.1. Methodological quality

The methodological quality of the included studies was generally high (mean global score±SD: 83.8%±14.2%), with scores ranging from 43.8%-100.0% (**Table 2**). Cohort and case studies (n=19) showed high scores for exposure measurement (criterion 5: 1.0±0.0), outcome assessment (criterion 10: 1.0±0.1), and statistical analysis (criterion 14: 0.9±0.2). High scores were also observed for identification of confounders (criterion 7: 0.9±0.3) and follow-up time reporting (criterion 11: 1.0±0.0, criterion 12: 1.0±0.0). Moderate scores were noted for the group similarity (criterion 2: 0.6±0.3), and confounding management strategies (criterion 8: 0.6±0.4)(**Figure 3**). Cross-sectional studies (n=29) showed high scores for inclusion and exposure criteria (criterion 1: 0.8±0.3; criterion 5: 1.0±0.0; criterion 6: 1.0±0.1), outcome assessment (criterion 10: 1.0±0.0), and statistical analysis (criterion 14: 0.9±0.3), with lower scores for confounding management strategies (criterion 8: 0.6±0.4)(**Figure 3**).

**Figure 3.**
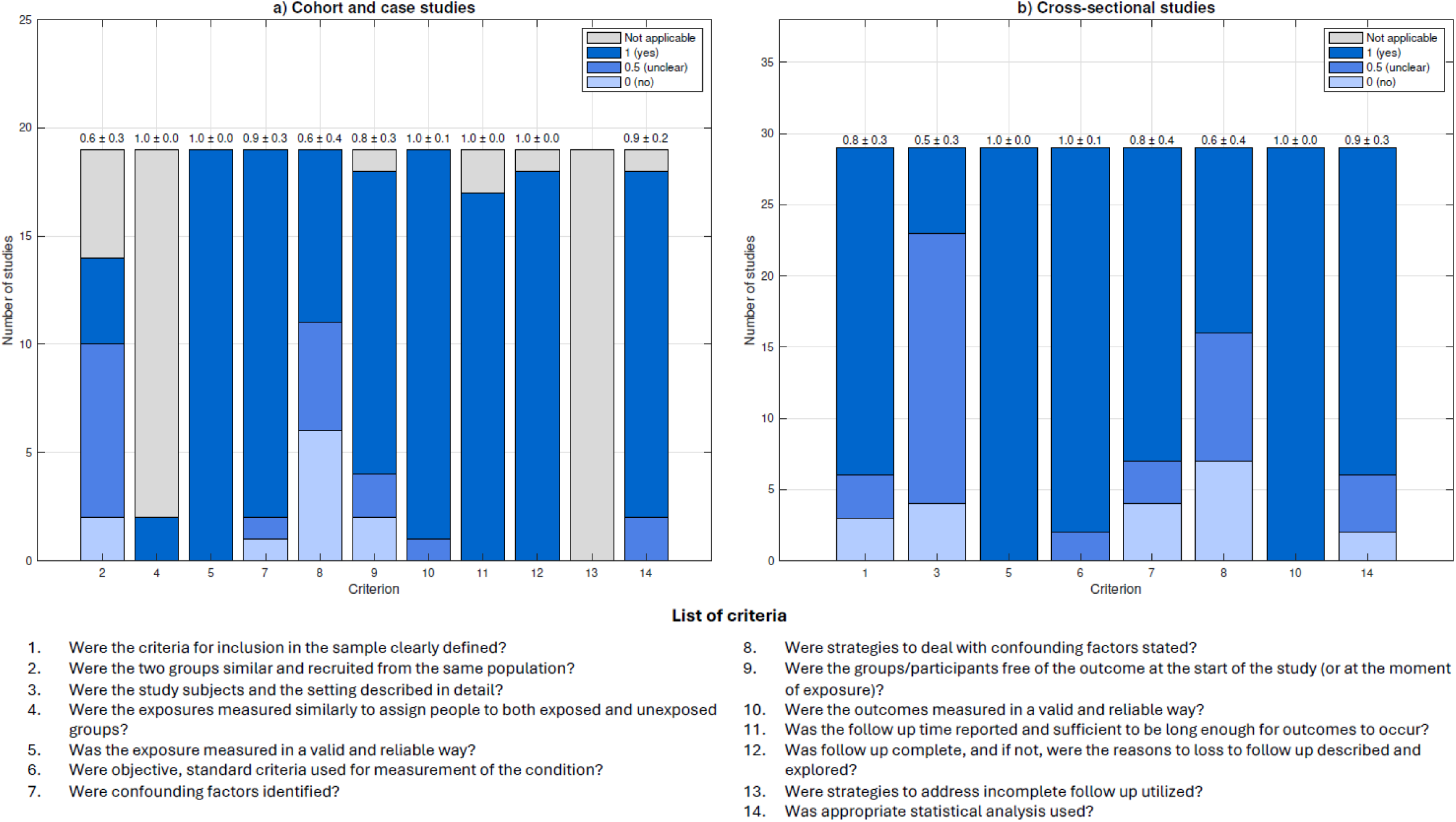
Quality assessment of included studies using the Joanna Briggs Institute critical appraisal checklists [21]. Stacked bar charts illustrate the distribution of scores assigned to each methodological criterion across (a) cohort studies and (b) cross-sectional studies. Each bar represents the number of studies that received a score of 0 (“no”), 0.5 (“unclear”), or 1 (“yes”) for a given criterion. The criteria are labeled numerically on the x-axis, and their full descriptions are listed below the figure. The mean score ± standard deviation is displayed for each criterion above each bar.

**Table 1.**
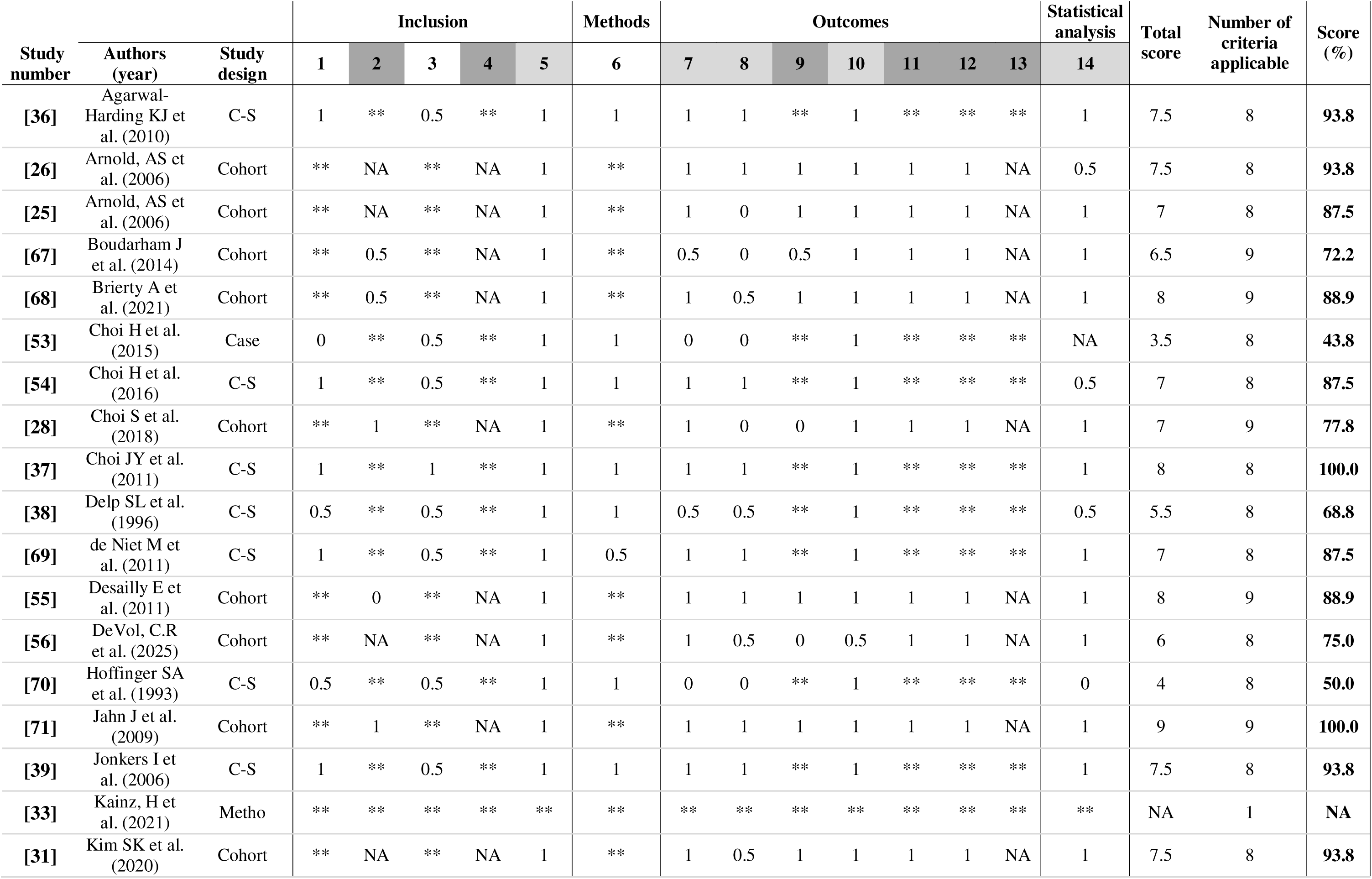
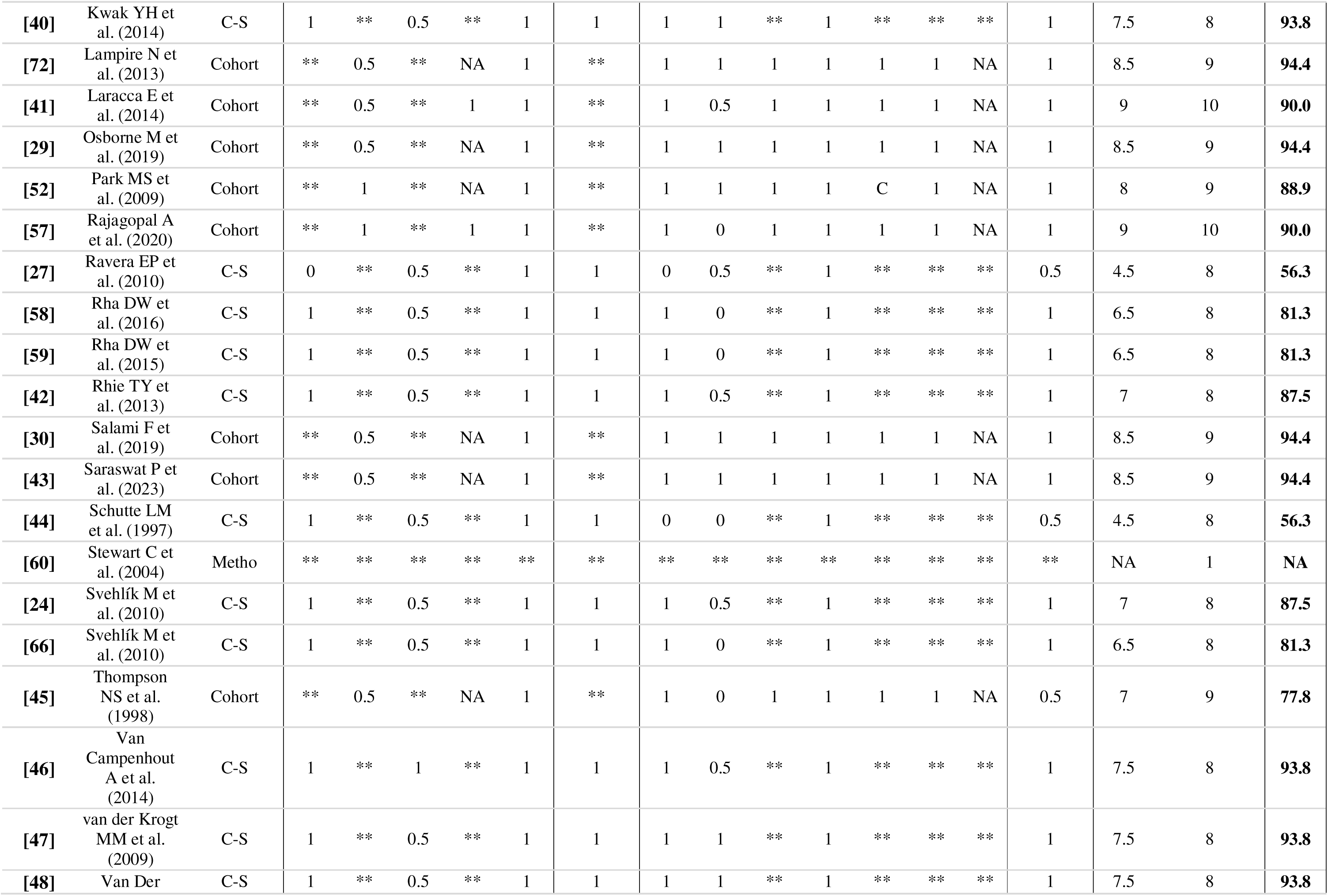
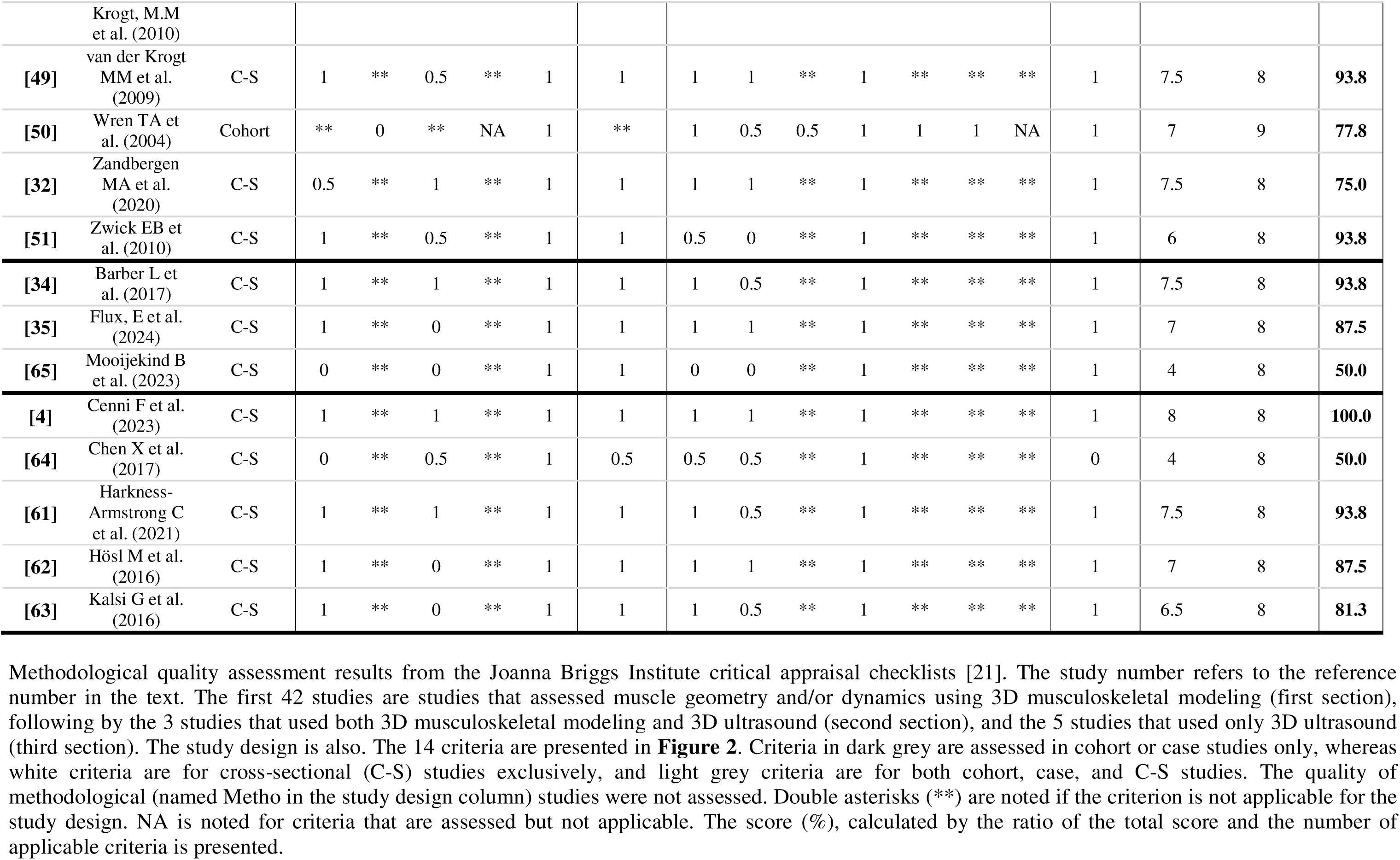
Methodological quality assessment results from the Joanna Briggs Institute critical appraisal checklists [21].

**Table 2.**
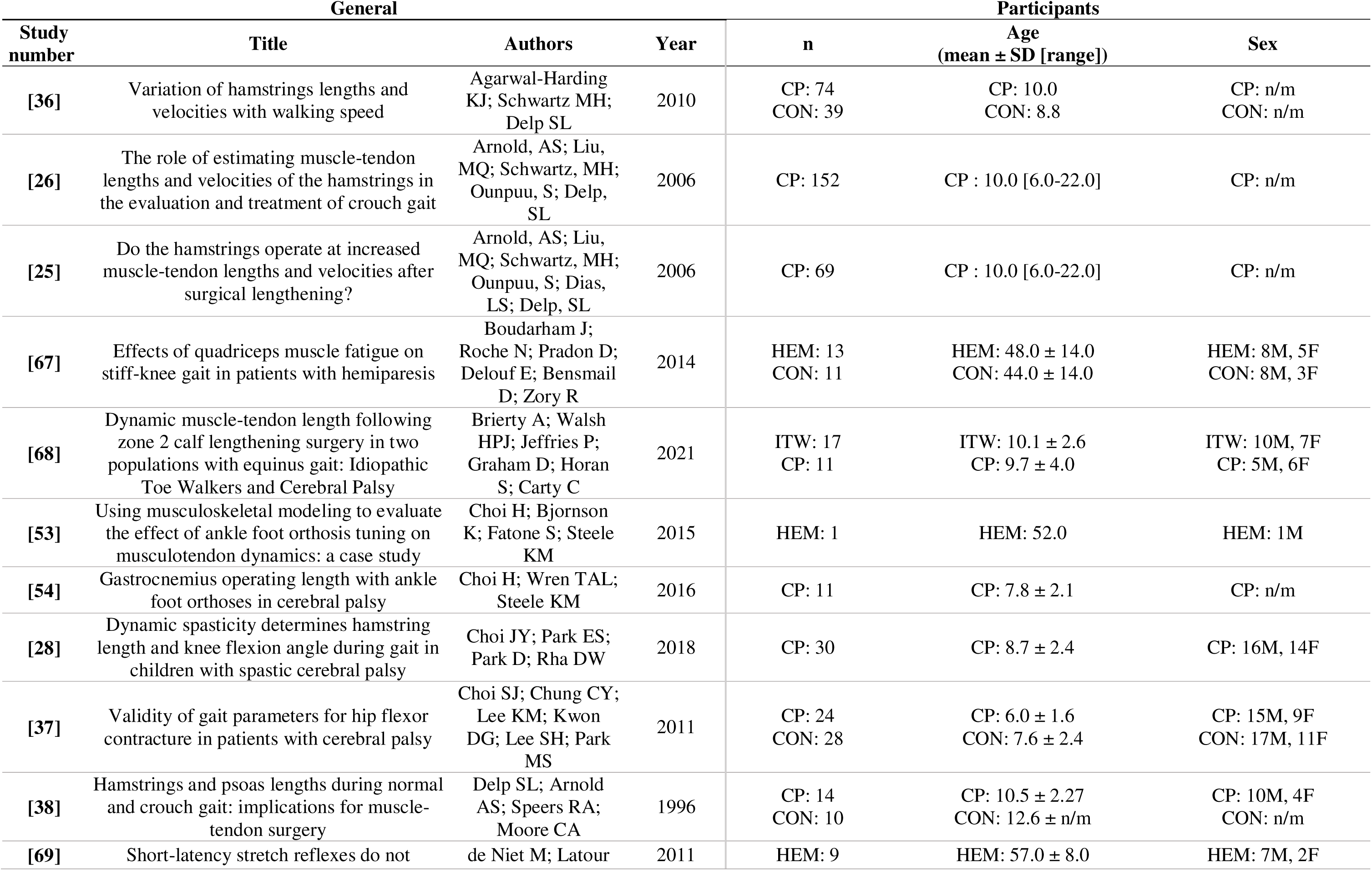
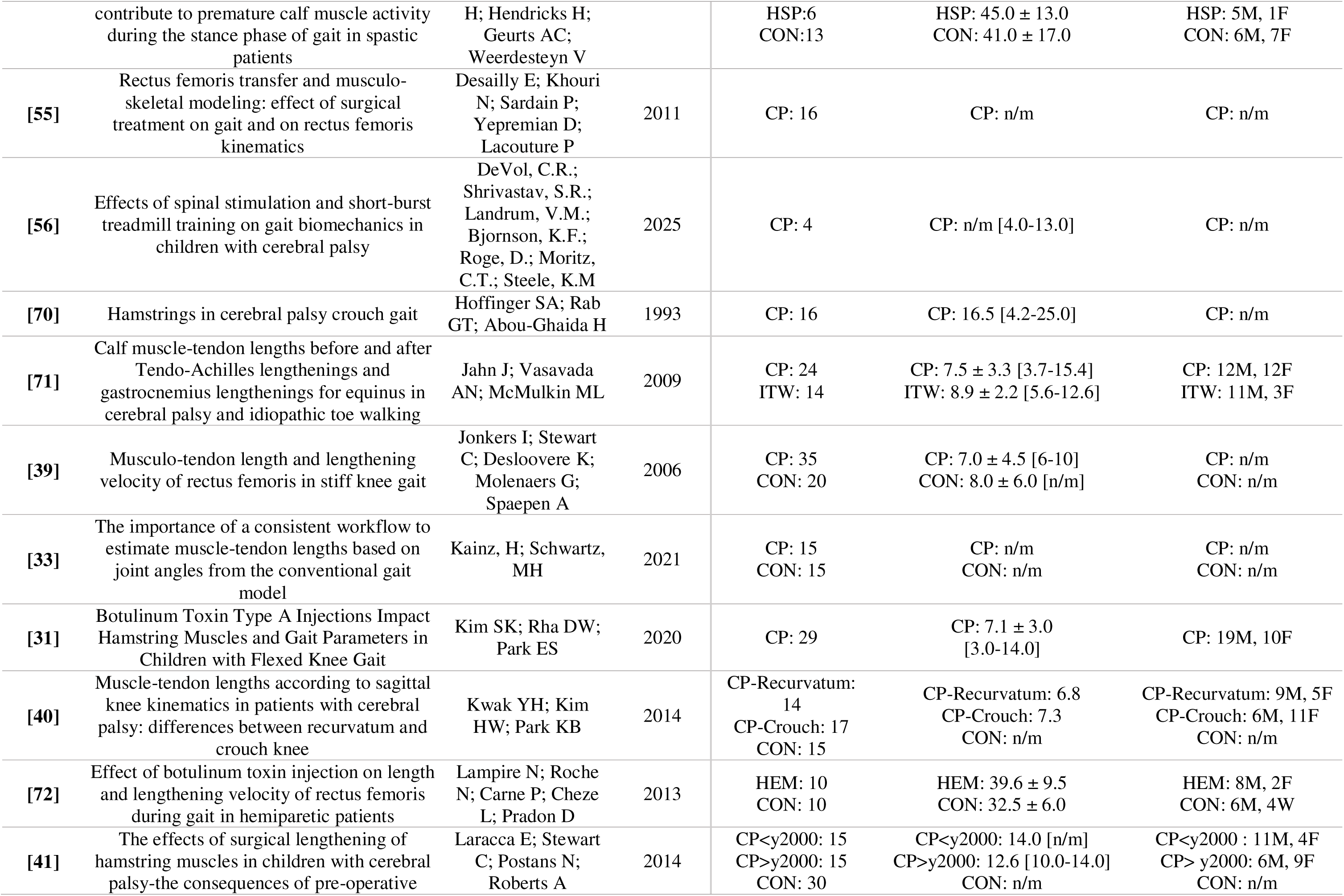
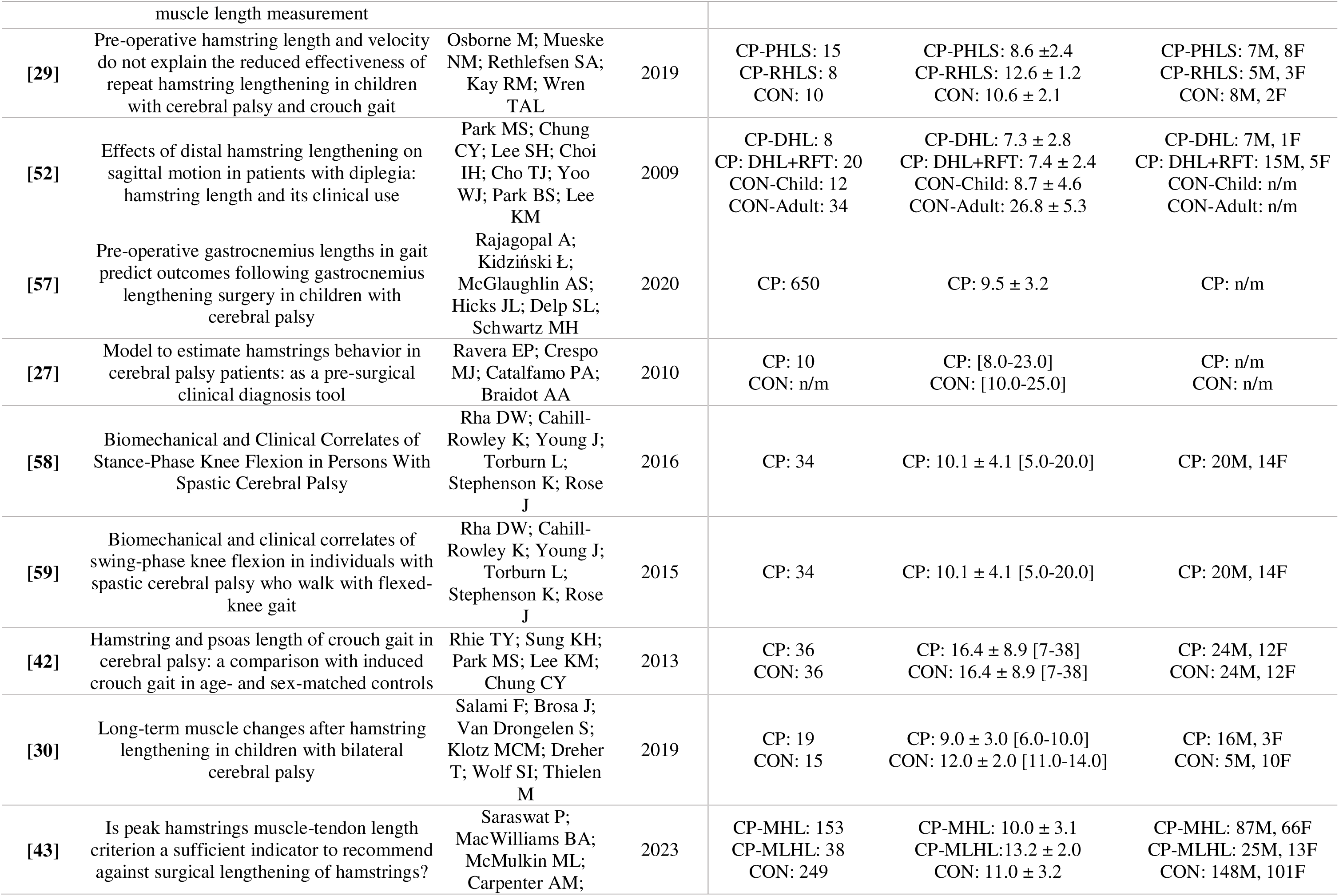
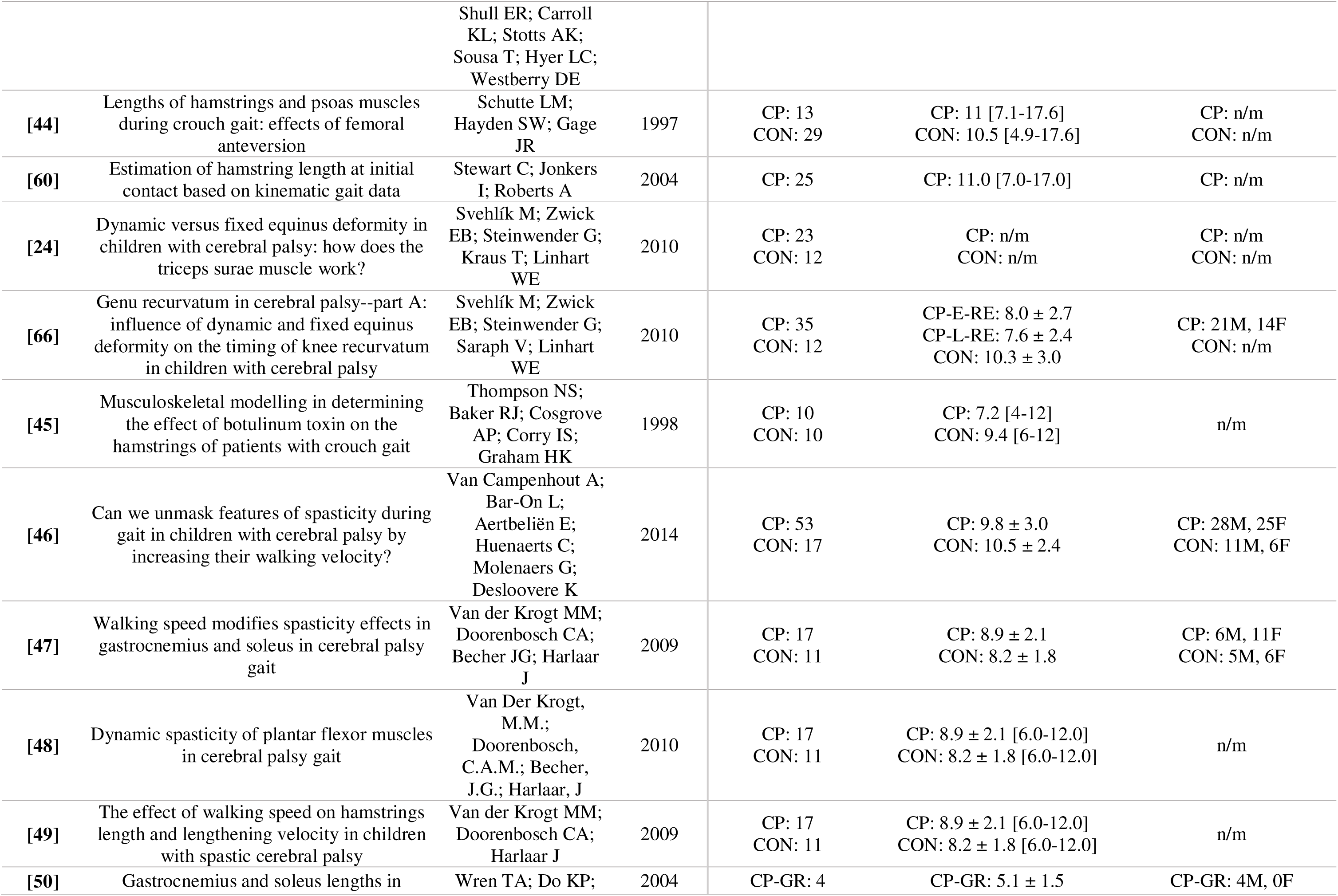
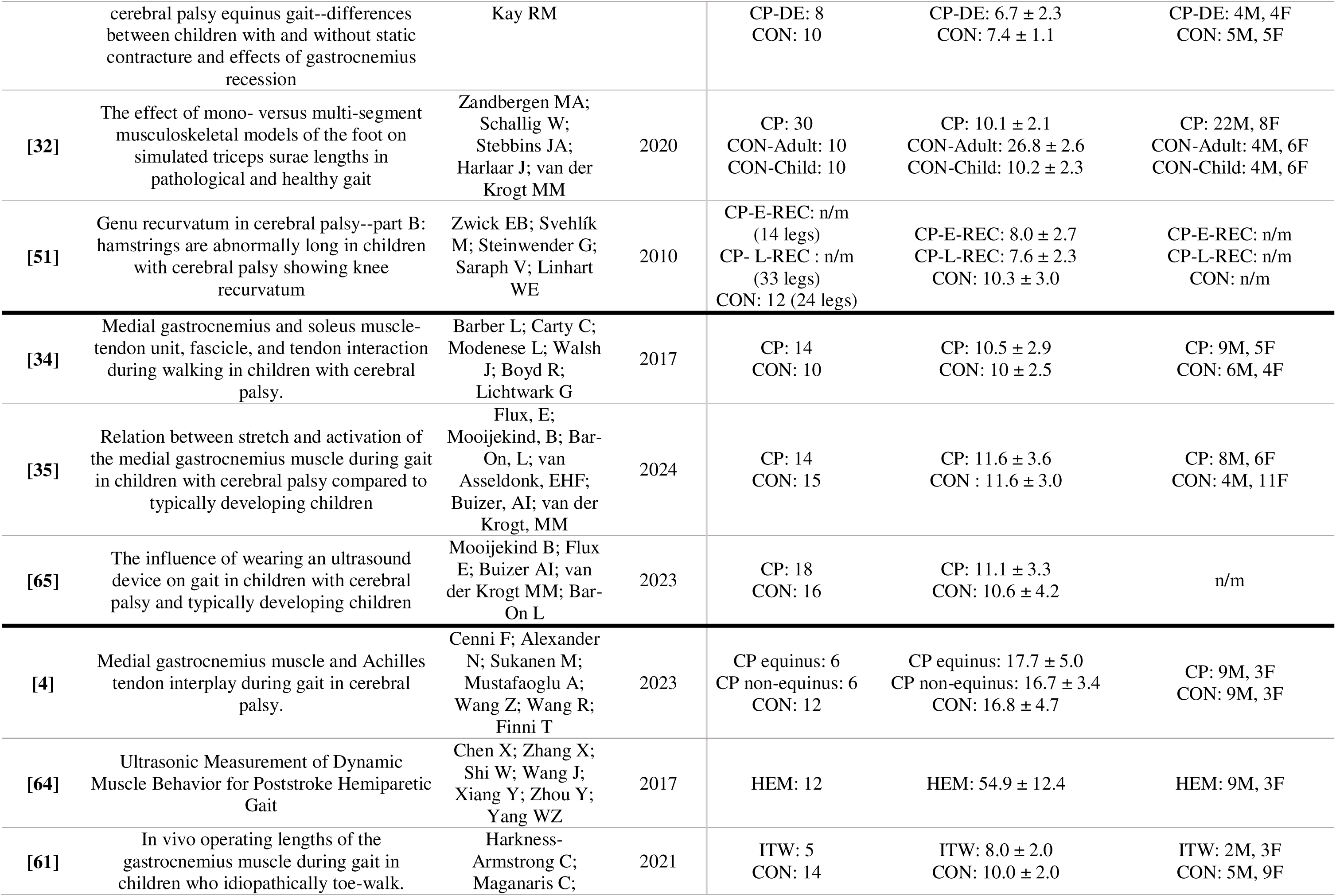
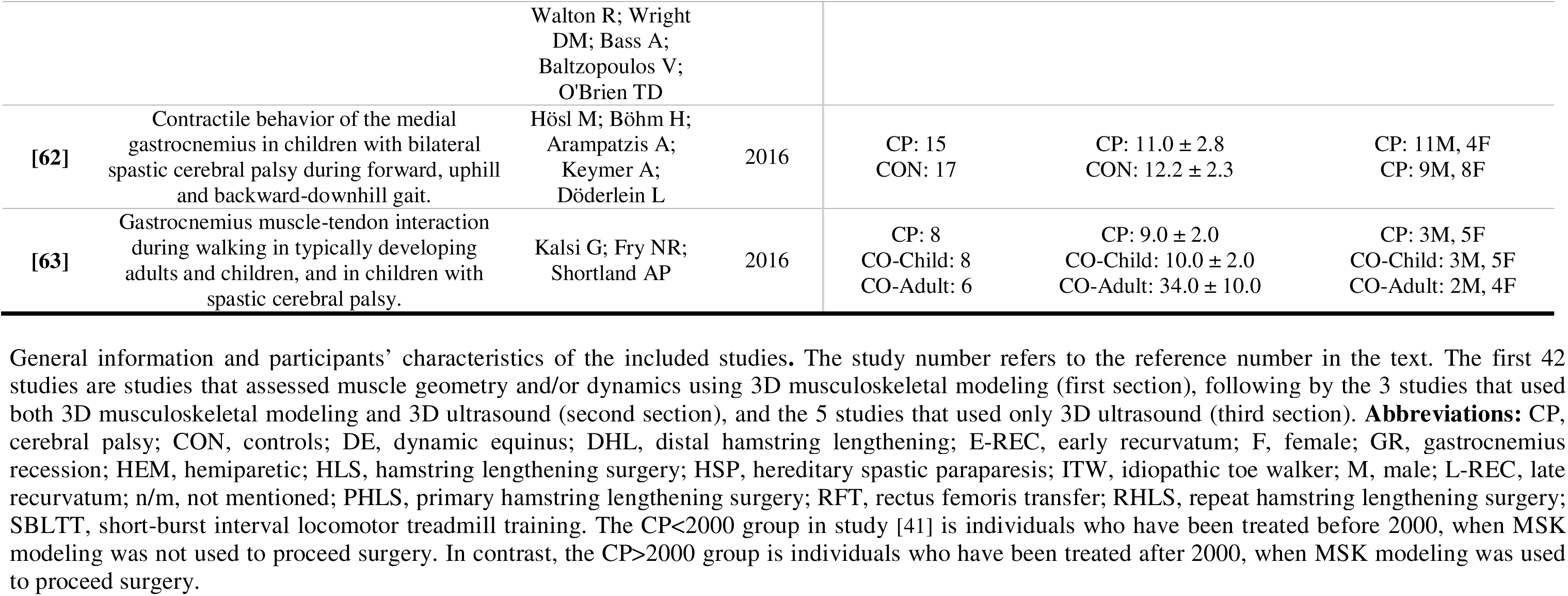
General information and participants’ characteristics of the included studies.

### 3.2. Musculoskeletal modeling

#### 3.2.1. Populations

Among the 45 studies that used MSK modeling, 27 compared children with CP to their control (CON) peers [24,27,29,30,32–52,65,66], 2 compared children with idiopathic toe walking (ITW) and children with CP [68,71], 3 compared adults with hemiparetic gait with their CON peers [67,69,72], and 12 focused on children with CP [25,26,28,31,54–60,70] or adults with hemiparetic gait [53] without comparing them with another population. Population characteristics are detailed for each study in **Table 2**. Both studies conducted by Arnold et al.(2006)[25,26], so as those of Van Der Krogt et al.(2009, 2010)[47–49] involve the same sample, given the similarity in population characteristics.

#### 3.2.2. Walking condition

A total of 2/45 studies assessed treadmill walking at self-selected speed (mean gait speed range = [0.7 - 1.0 m/s])[35,65], whereas 43/45 studies focused on overground walking in a laboratory setting [4,24–34,36–60,66,67,69–72]. Among them, 26/43 were conducted at self-selected comfortable speed [4,24–26,28–32,34,39–41,43,44,51,54,56–59,66,67,69,71,72], 4/43 at different walking speed (e.g., fast, slow, comfortable speed)[36,47–49], and 13/43 studies did not mention the speed condition [27,33,37,38,41,42,45,50,52,54,55,60,70]. Participants were walking barefoot in 17/45 [24–26,28,29,31,32,34,37,40,46,52,56,58,59,68,72], shod in 3/45 [35,53,65], barefoot then shod in one [54], and 24/45 did not mention the footwear condition [27,30,33,36,38,39,41–45,47–51,55,57,60,66,67,69–71]. Methodological details of each study are provided in **Table 3**.

**Table 3.**
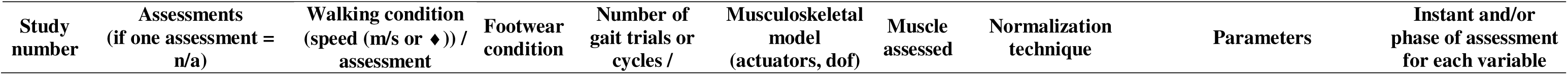
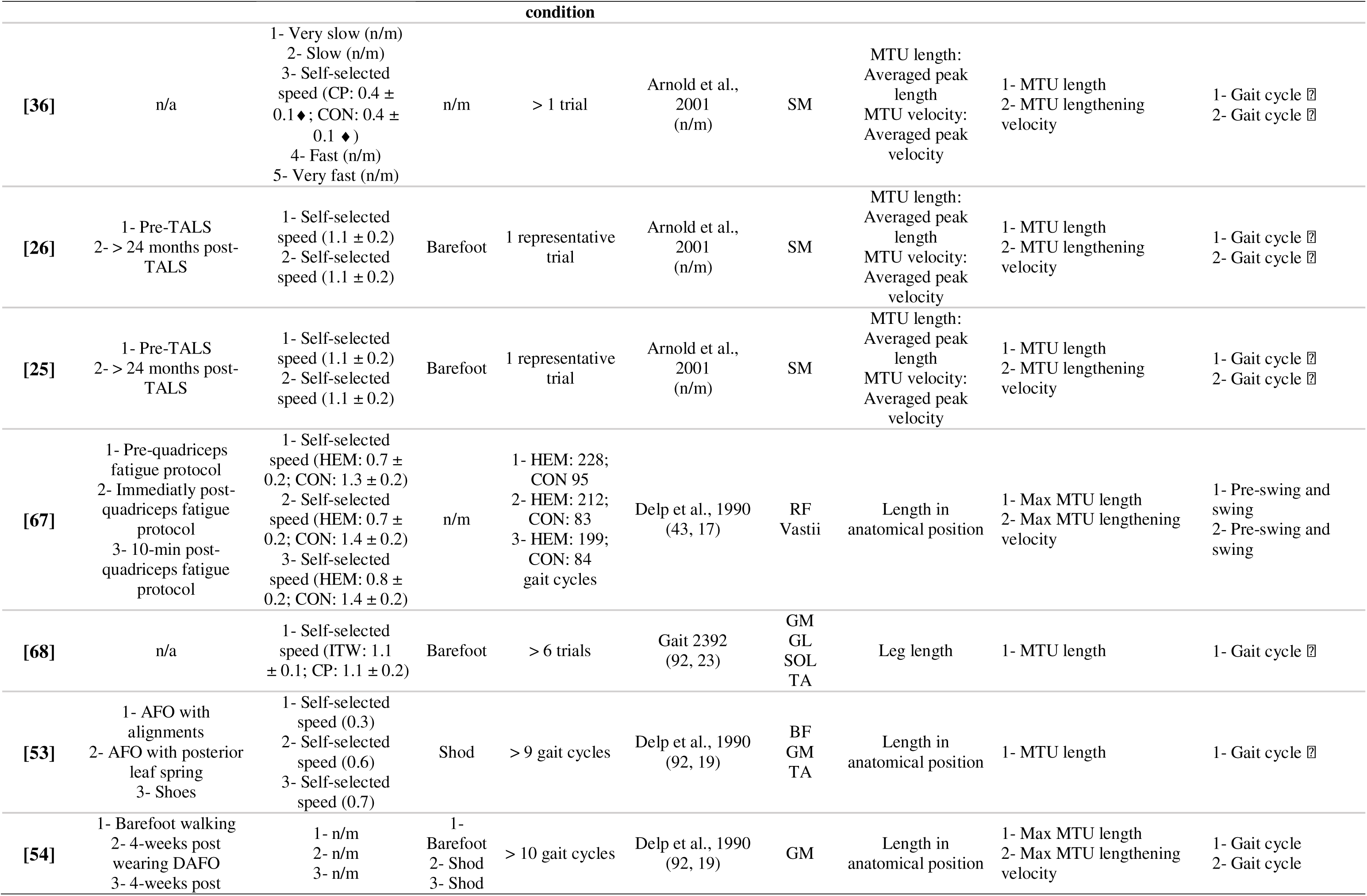
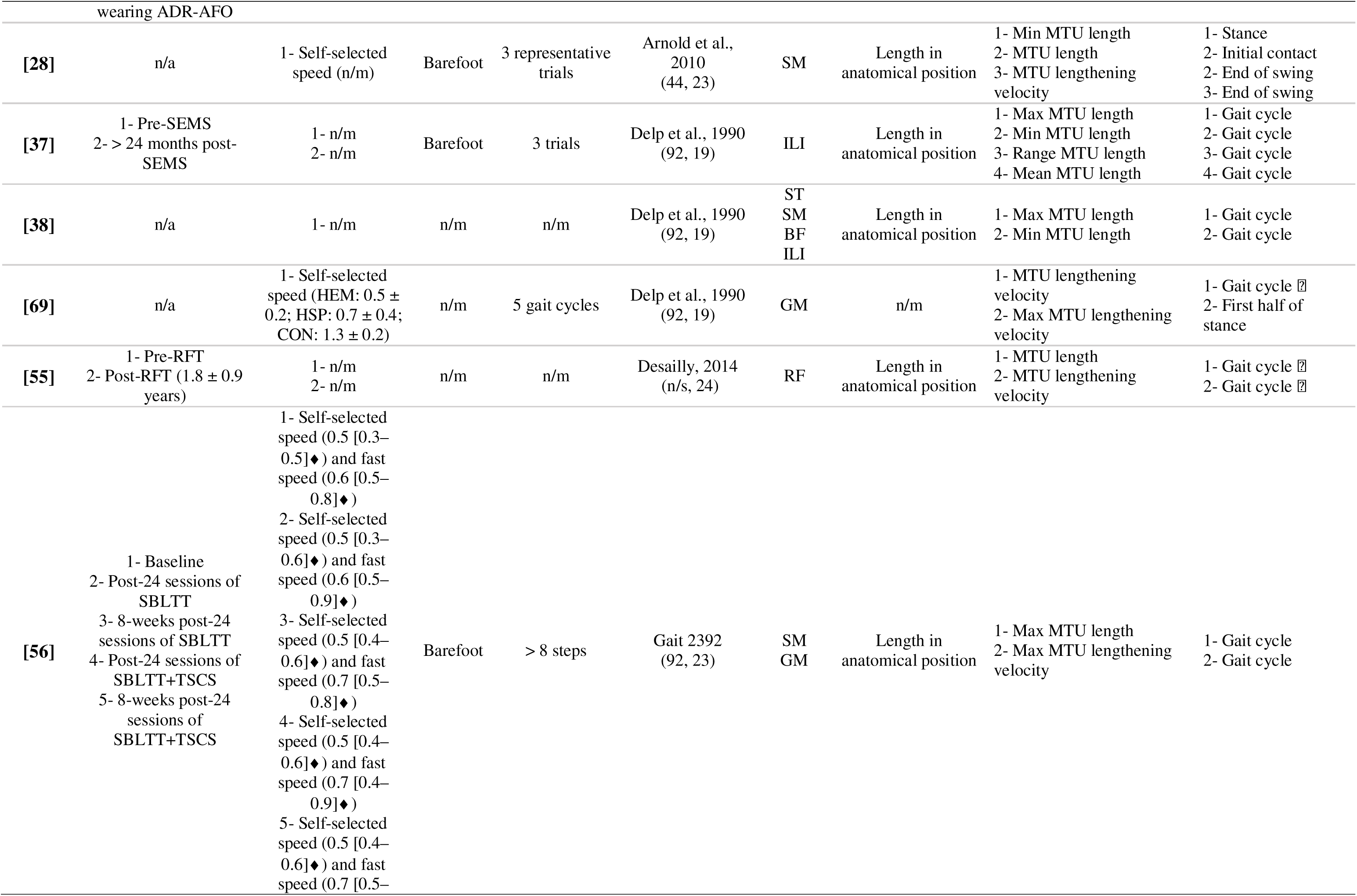
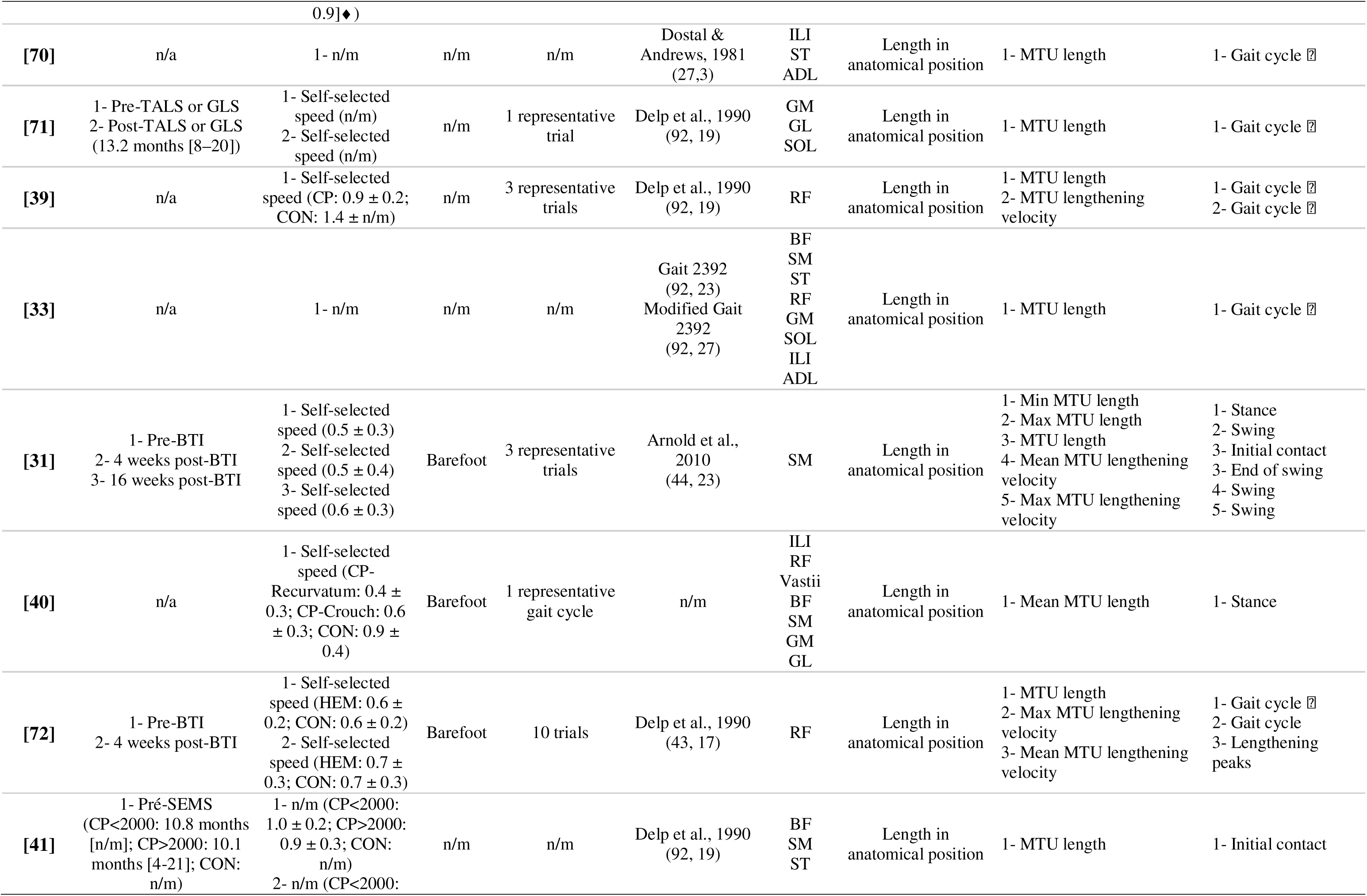
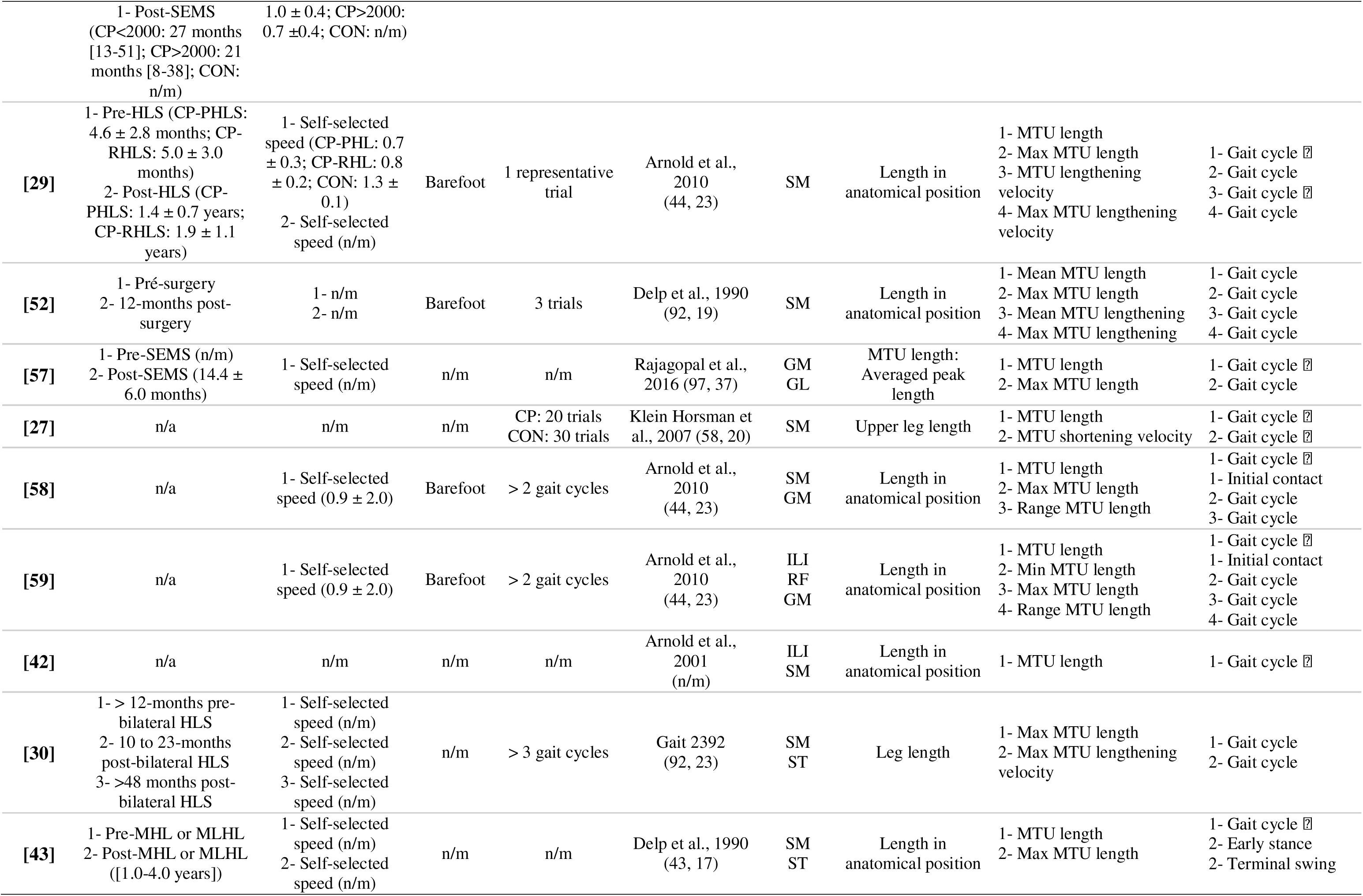
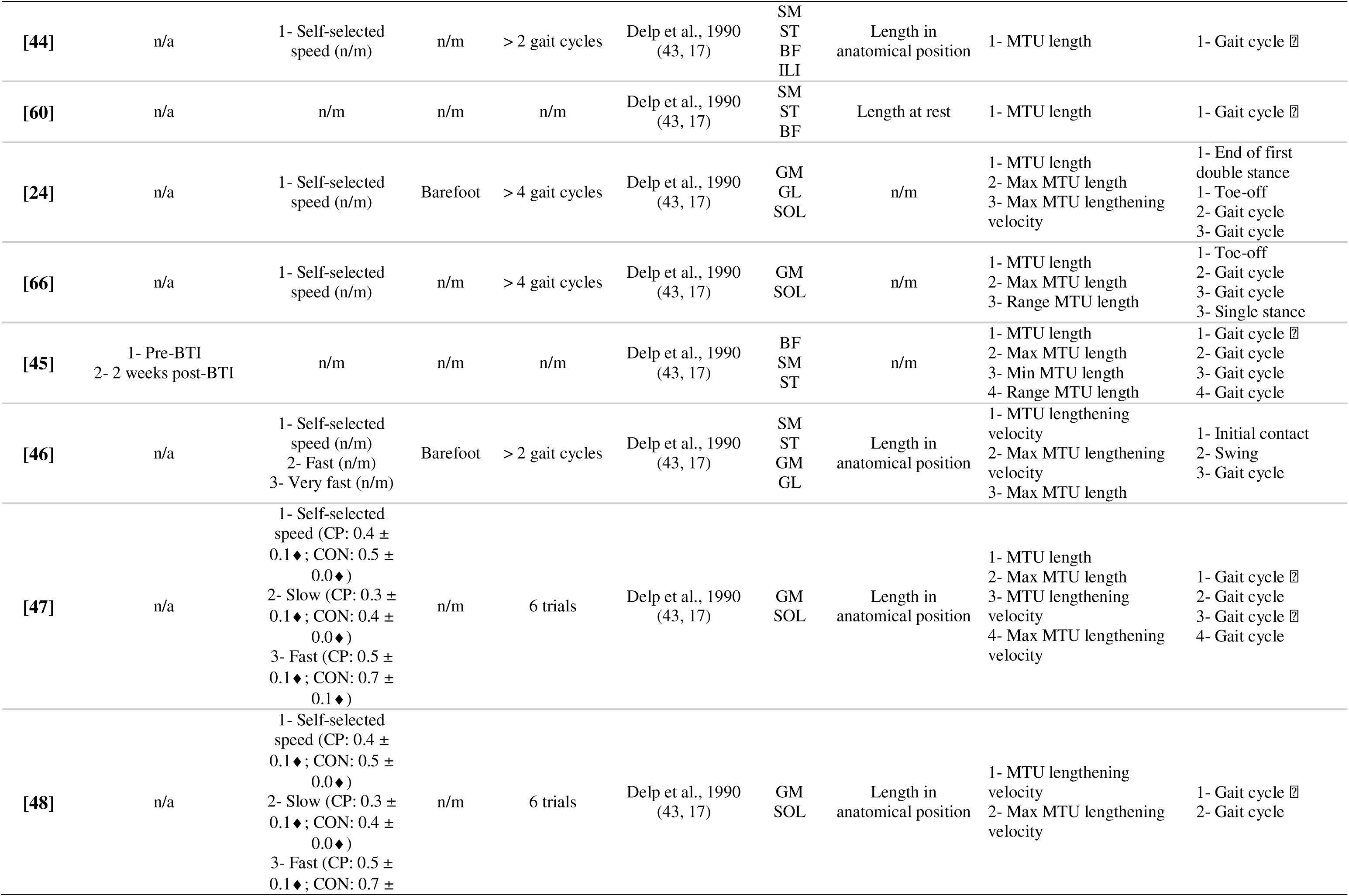
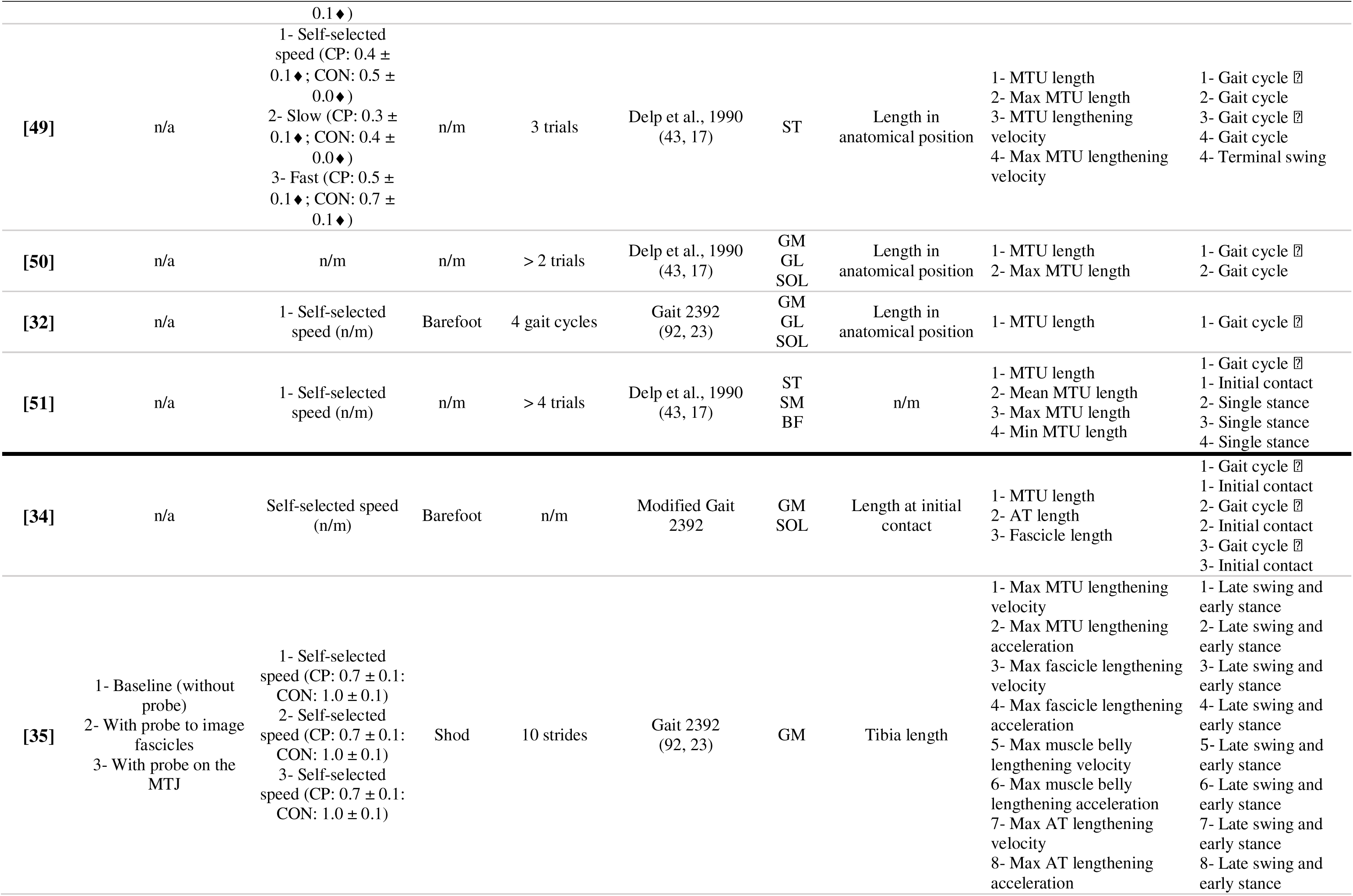
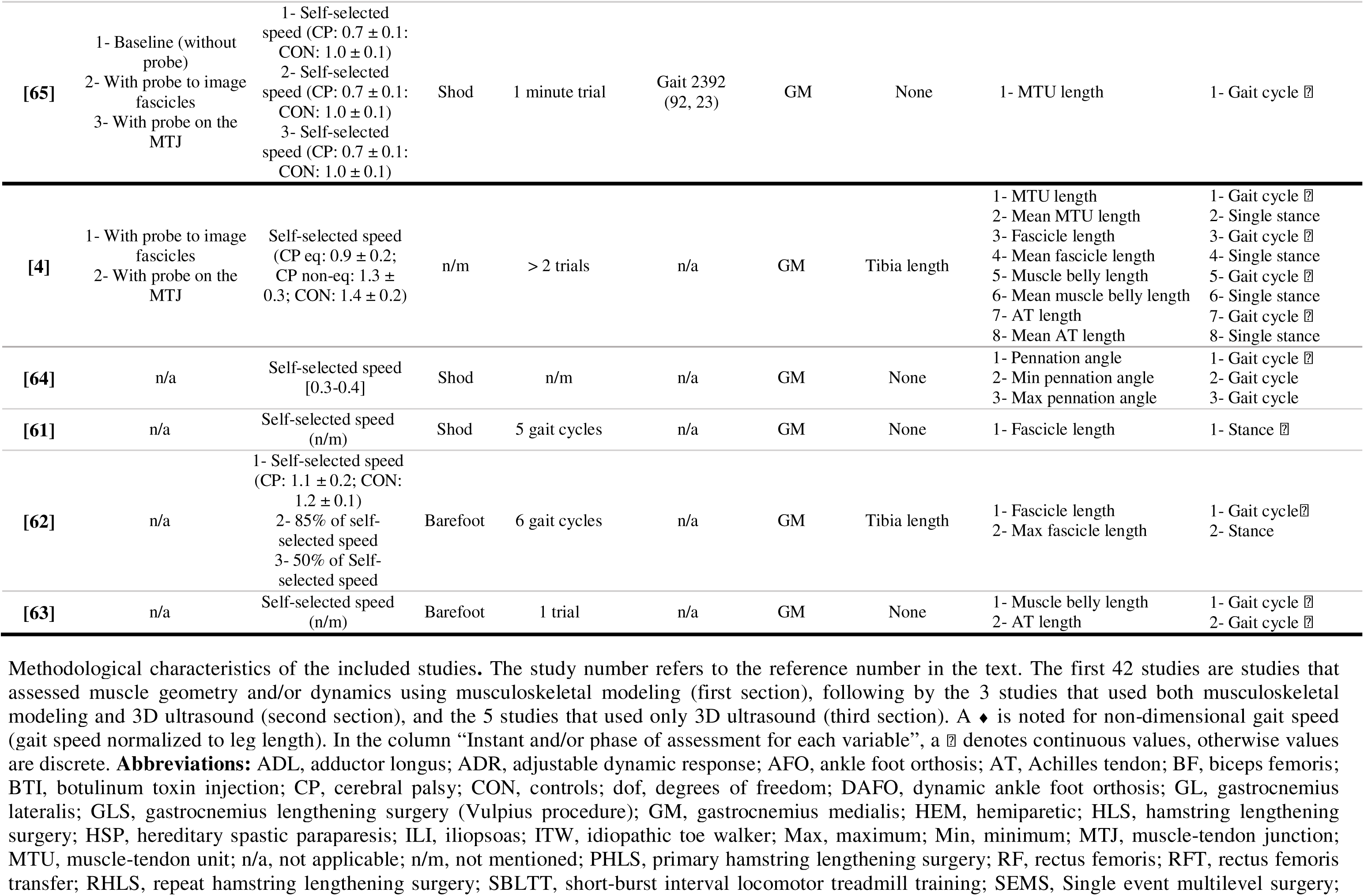
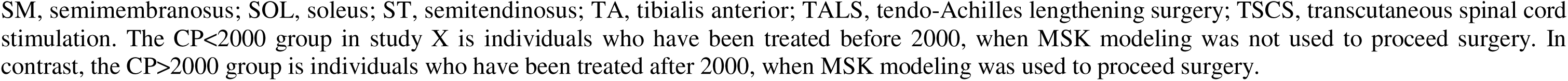
Methodological characteristics of the included studies.

#### 3.2.3. Kinematic and musculoskeletal models

All studies were tracking retro-reflective marker trajectories using 8-12-cameras optoelectronic motion capture system. For computing gait kinematics, 36/45 studies used the Conventional Gait Model (CGM)[24–34,36–38,40–46,50–55,57,59,60,66–69,71,72] or an alternative [73], whereas 2/45 used the Human Body Model (Motek Medical, Amsterdam, Netherlands), and 3/45 used the CAST model [47–49]. Regarding the musculoskeletal model, 8 different models were used across studies. Among these, 6 were OpenSim-based: The Gait2392 model or a modified version [11](n=8/45 studies [30,32–35,56,65,68]), the Arnold et al.(2001) model [74](n=4/45 studies [25,26,36,42]), the Arnold et al.(2010) model [8] (n=5/45 studies [28,29,31,58,59]), the Delp et al.(1990) model [75](n=23/45 studies [24,37–39,41,43–51,51–54,60,67,69,71,72]), the Klein Horsman et al.(2007) model [76](n=1/45 studies [27]), the Rajagopal et al.(2016) model [77](n=1/45 studies [57]). Two studies used non-OpenSim-based models: The Desailly (2008) model [78](n=1/45 studies [55]) and the Dostal & Andrew (1981) model [79](n=1/45 studies [70]). One study did not specify which MSK model was used [40]. The number of degrees of freedom and actuators for each model is detailed in **Table 3**. All included studies used scaled generic MSK models, where joint axes and segment lengths were adjusted based on anatomical markers. Only 3 studies reported subject-specific personalization [44,55,60]. All of them personalized the femoral anteversion by creating an additional joint into the intertrochanteric region of the femur [60], or by using a deformed model representing the femoral anteversion [44,55]. One of them also personalized the axial torsion of the tibia based on the clinical evaluation [55].

#### 3.2.4. Outcomes

A total of 11 muscles were assessed across studies: semimembranosus (n=22/45 studies [25–31,33,36,38,40–46,51,52,56,58,60]), gastrocnemius medialis (n=21/45 studies [24,32–35,40,46–48,50,53,54,56–59,65,66,68,69,71]), semitendinosus (n=12/45 studies [26,30,33,41,43–46,49,51,60,70]), soleus (n=10/45 studies [24,32–34,47,49,50,66,68,71]), biceps femoris (n=9/45 studies [33,38,40,41,44,45,51,53,60]), gastrocnemius lateralis (n=8/45 studies [24,32,40,46,50,57,68,71]), iliopsoas (n=8/45 studies [33,37,38,40,42,44,59,70]), rectus femoris (7/45 studies [33,39,40,55,59,67,72]), tibialis anterior (n=2/45 studies [33,53]), vastii (n=2/45 studies [40,67]) and adductor longus (n=1/45 studies [33]). All but 2 studies [48,69] assessed the MTU length during gait, and 20/45 studies assessed the MTU lengthening velocity [24–31,35,36,46–49,54–56,67,69,72]. Only one study assessed MTU lengthening acceleration [35].

A body chart representing the reported outcomes for each muscle is presented in **Figure 4**. Most studies (42/45 studies) reported the MTU length during gait. Among them, 27 [25–27,29,32–34,36,39,42–45,47,49–51,53,55,57–60,65,68,71,72] reported the MTU length over the full gait cycle (continuous values), while 8 studies provided MTU length values at specific instants (discrete values): at initial contact [28,34,41,51,58], at the end of first double stance [24], at toe-off [66], and at the end of the swing phase [28], or mean values during a specific gait phase: during single stance [40]. Regarding other discrete values, the minimal MTU length during stance phase was reported in 7 studies [28,31,37,38,45,51,59], and the maximal MTU length was reported in 22 studies: during the swing phase in 3 studies [31,43,67], during pre-swing in 1 study [67], at initial contact in 1 study [43], during single stance in 1 study [51], and as the peak value during the gait cycle 16 studies [24,25,29,30,37,45–47,49,50,52,54,56–59]. Regarding MTU lengthening velocity, the value was reported over the full gait cycle in 9 studies [25–27,36,39,47,49,55,69], whereas other studies reported discrete values at the end of the swing phase [28], at initial contact [46], or averaged across the entire gait cycle [29], the swing phase [31], or at lengthening peaks [72]. The maximal MTU lengthening velocity was reported as the peak value during the gait cycle in 7 studies [24,30,47–49,54,56], during swing phase in 3 studies [31,46,67], during the pre-swing phase in 1 study [67], during the first half of the stance phase in 1 study [69], or during late swing and early stance in 1 study [35]. The maximal MTU lengthening acceleration was reported as the peak value during late swing and early stance in 1 study [35].

**Figure 4.**
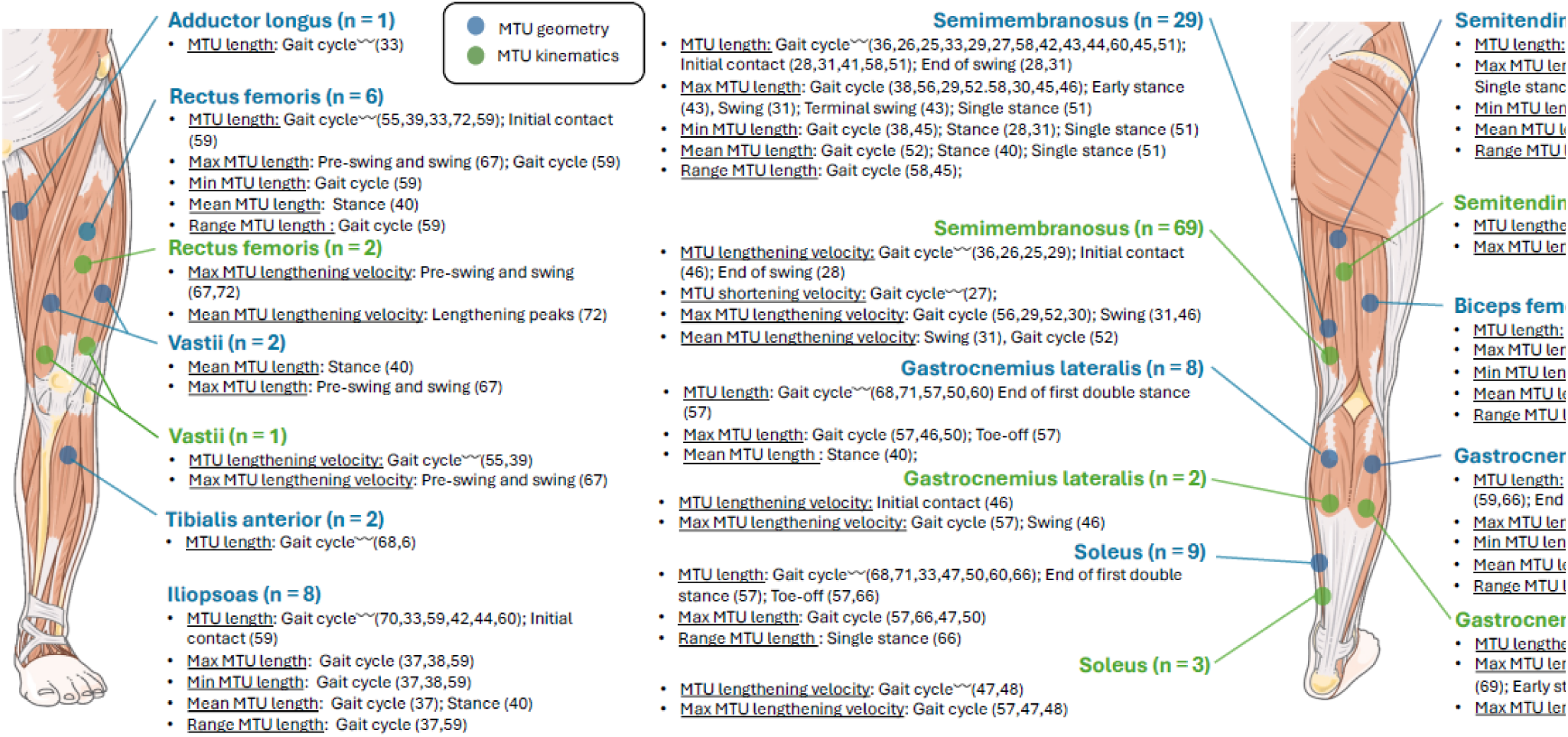
Each muscle assessed is presented, with the number of studies over the 45 included studies using musculoskeletal modeling in parentheses. For each muscle, the parameters and the instant/moment of assessment are indicated (〰□ denotes continuous values, otherwise values are discrete), followed by the study references in parentheses. The iliopsoas and the vastus intermedius (from the vastii) are not linked to a specific body segment in the schematic illustration, as its deep anatomical location prevents reliable surface representation.

Among the 45 studies that reported the MTU length, 29 normalized it to the muscle length in anatomical position (i.e., all joints positioned at 0° of flexion)[28,29,31–33,37–44,46–50,52–56,58,59,67,70–72](biceps femoris, semimembranosus, semitendinosus, rectus femoris, vastii, gastrocnemius medialis, gastrocnemius lateralis, soleus, iliopsoas, adductor longus), 4 to the average peak length [25,26,36,57](semimembranosus, gastrocnemius medialis, gastrocnemius lateralis), 2 to the leg length [30,68](semimembranosus, semitendinosus, gastrocnemius medialis, gastrocnemius lateralis, soleus, tibialis anterior)(gastrocnemius medialis), 1 to the upper leg length [27](semimembranosus), 1 to the length at initial contact [34] (gastrocnemius medialis, soleus), 1 to the length at rest [60], and 6 did not normalize or mention the normalization technique [24,24,45,65,66,69](gastrocnemius medialis, gastrocnemius lateralis, soleus, biceps femoris, semimembranosus, semitendinosus). Regarding MTU lengthening velocity, 2 studies normalized it to the averaged peak velocity [25,26] and 1 to the tibia length [35]. For the only study that assessed the MTU lengthening acceleration, it was normalized to the tibia length [35].

#### 3.2.5. Studies’ aims and design

A total of 25/45 studies followed a cross-sectional study design [24,27,28,32,34–36,38–40,42,44,46–51,53,58,59,65,66,69,70], focusing on observation and/or group comparisons at a single point in time. Moreover, 18/45 studies used a cohort (n=17)[25,26,29–31,37,41,43,45,52,55–57,67,68,71,72] or case (n=1)[53] study design involving observations and/or group comparisons before and after an intervention. Finally, 2/45 studies used a methodological study design [33,60].

Among the 25 cross-sectional studies, 9 used MTU length either as a classification criterion for patients [24,27,66], as an explanatory factor for gait deviations [51,58,59], or in relation to other variables such as muscle activity and spasticity [28,48,69]. A total of 7 studies assessed MTU length or lengthening velocity across different gait patterns (e.g., crouch gait, stiff knee gait)[38–40,42,50,70] or different conditions (e.g., with or without orthoses)[54], while 5 examined their relationship with walking speed [36,46,47,49] or muscle activation [35]. Also, 2 studies evaluated validity of MSK modeling in case of femoral [44] or foot [32] deformities, 1 study explored the impact of wearing an ultrasound probe on gait parameters [65], and 1 study the muscle-tendon interactions during gait [34].

Among the 18 cohort studies, 13 investigated the effects of therapeutic interventions on muscle length and/or lengthening velocity: 8 focused on surgical procedures [25,29,30,41,52,55,68,71], 3 on botulinum toxin injections [31,45,72], and 2 on fatigue or rehabilitation protocols [56,67]. A total of 5 studies examined whether muscle length and/or lengthening velocity could serve as relevant clinical parameters [37] or predictors of surgical outcomes [26,43,52,57].

The 2 methodological studies explored different approaches to improve muscle length estimations [33,60], whereas the case study examined the effects of different types of orthoses on muscle length during gait [53].

### 3.3. Ultrasound imaging

#### 3.3.1. Populations

Among the 8 studies that used USI, 6 compared CP to CON children [4,34,35,62,63,65] or to CON adults [63],1 focused on post-stroke survivors [64], and 1 examined children with ITW [64]. Population characteristics are detailed for each study in **Table 2**.

#### 3.3.2. Walking condition

A total of 5/8 studies assessed treadmill walking at self-selected speed (mean gait speed range = [0.3–1.2 m/s])[35,61,62,64,65], while 3/8 focused on overground walking in a laboratory setting at comfortable self-selected speed (mean gait speed range = [0.9–1.4 m/s])[4,34,63]. Participants were walking barefoot in 3 studies [34,62,63], shod in 4 studies [35,61,64,65], and 1 study did not mention the footwear condition [4]. Methodological information of each study is provided in **Table 3**.

#### 3.3.3. Kinematics and ultrasound devices

Among the 8 studies, 7 tracked markers displacement to calculate joint kinematics using 8-12-cameras optoelectronic system, while 1 used an electronic goniometer (ankle kinematics)[64]. A 59 [35,65] or 60 mm [4,34,61–63] linear probe was used in all but one [64] study, that did not mention probe size. Also, 1 study did not mention how USI data was processed [34] and another did not process USI data [65].

Py3DFreehandUS, an algorithm developed by Cenni et al.(2020)[80], was used by 2 studies to track and calculate the MTJ displacement [4,35], whereas 1 used manual tracking [63]. To track fascicles and calculate fascicle length during gait using automatic algorithms, 2 studies [35,61] used ImageJ [81], 1 study [4] used UltraTrack [82], and 1 [62] used the algorithm developed by Gillet et al.(2013)[83]. To track pennation angle during gait, one study used a Gabor Filter algorithm [64].

#### 3.3.4. Outcomes

Of the 8 studies included, 7 focused exclusively on the gastrocnemius medialis [4,35,61–65] and 1 investigated both gastrocnemius medialis and soleus [34].

A body chart representing the reported outcomes for each muscle is presented in **Figure 5**. Among the 8 studies, only 1 reported the MTU length over the full gait cycle (continuous values) and the average across single stance (discrete values)[4]. Half of the studies (4/8 studies) measured fascicle length of the gastrocnemius medialis [4,34,61,62] or of the soleus [34]. The fascicle length was reported over the full gait cycle in 3 studies [4,34,62], over the stance phase in 1 study [61] or averaged during single stance in 1 study [4], and at initial contact in 1 study (discrete value)[34]. The maximum value during stance (discrete value) was also reported in 1 study [62]. The Achilles tendon (AT) length was reported over the full gait cycle in 3 studies, for the gastrocnemius [4,34,63] and the soleus [34] parts of the AT. One study reported the average AT length of the gastrocnemius part during single stance [4], and another reported the length of the gastrocnemius and the soleus parts at initial contact [34]. Regarding the muscle belly length, it was reported over the full gait cycle by 2 studies [4,63] and averaged during single stance in 1 study [4]. Only 1 study calculated the pennation angle and reported it over the full gait cycle [64], as well as the minimum and maximum values. Finally, 1 study calculated AT, muscle belly and fascicle lengthening velocity and acceleration [35], and reported the maximal value during late swing and early stance.

**Figure 5.**
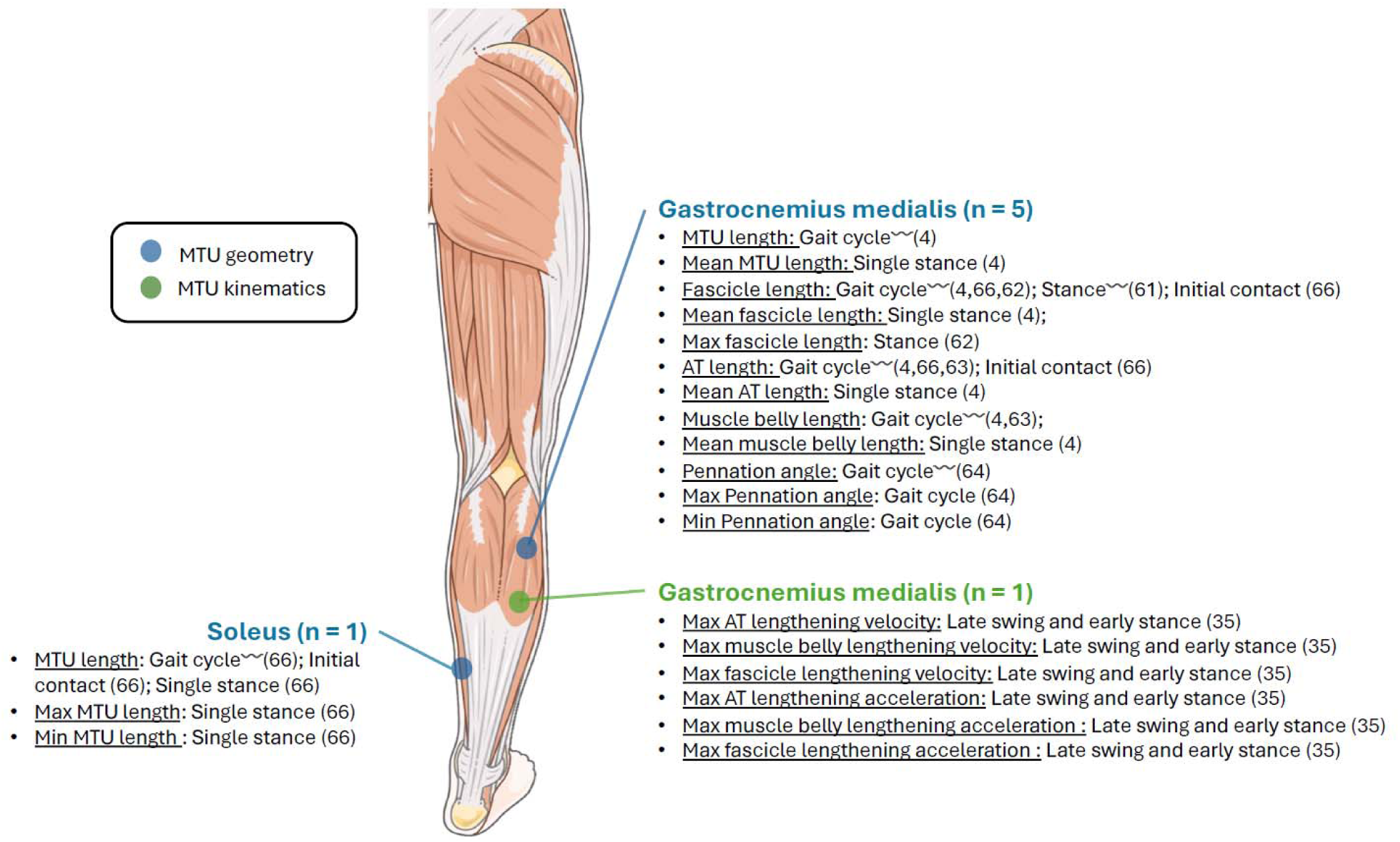
Each muscle assessed is presented, with the number of studies over the 8 included studies using 3D ultrasound in parentheses. For each muscle, the parameters and the instant/moment of assessment are indicated (〰□ denotes continuous values, otherwise values are discrete), followed by the study references in parentheses.

The MTU length was calculated as the sum of the Euclidean distance from the center of the knee joint to the gastrocnemius MTJ and the Euclidean distance from the gastrocnemius MTJ to the marker on the calcaneus [4]. Fascicle length was measured by placing an ultrasound probe over the belly of the gastrocnemius [4,34,61,62] or the soleus [34]. The AT length was calculated by securing an ultrasound probe over the MTJ and by following its displacement in 2 studies [4,63]. AT length was then calculated as the distance between MTJ and calcaneus marker. The other study calculated AT length (both gastrocnemius and soleus parts) by subtracting muscle belly length estimated by fascicle length multiplied by the cosinus of the pennation angle, to the MTU length (as calculated by Fukunaga et al. [84])[34]. The muscle belly length was reported as the distance between MTJ and knee joint center [4] or by subtracting AT length (calculated as the distance between MTJ and insertion point of the tendon) to MTU length (derived from the imaging-based model of Eames et al. [10]). The fascicle lengthening velocity was calculated by differentiating the fascicle length, which was obtained by securing an ultrasound probe over muscle belly. The muscle belly lengthening velocity was calculated by differentiating the muscle belly length, calculated as the distance between the MTJ position (estimated by securing an ultrasound probe over the MTJ) and the virtual origin of the GM muscle. Finally, the AT lengthening velocity was calculated by differentiating the AT length, calculated as the distance between the MTJ to the calcaneus marker. All acceleration metrics were calculated by differentiating the lengthening velocity values.

The only study that assessed the MTU length normalized the measurement to the tibia length [4]. Among the 4 studies that reported the fascicle length, 2 normalized it to the tibia length [4,62], 1 to the length at initial contact [34], and 1 did not normalize [61]. Regarding the 2 studies that assessed muscle belly length, 1 normalized it to tibia length [4] whereas the other did not normalize it [63]. For the 3 studies that assessed AT length, the parameter was normalized to the length at initial contact [34], to tibia length [4], or was not normalized [63]. The pennation angle was not normalized [64]. Finally, MTU, AT, muscle belly and fascicle lengthening velocity and acceleration were normalized to the tibia length [35]

#### 3.3.5. Studies’ aims and design

All 8 studies used a cross-sectional design, making observations at a single point in time. 7 of them focused on comparison between control and populations with neuromotor impairments [4,34,35,61–63,65], while 1 focused on comparisons between the affected and the unaffected side of stroke populations [64]. Among them, 5 were observational studies that aimed at reporting length-related values (MTU, fascicle, and/or tendon)[4,34,62,63], or the pennation angle [64] during gait, 1 examined the impact of wearing an ultrasound probe during gait on joint kinematics and MTU length [65], 1 assessed the relationship between muscle activation and peak values of tendon, fascicle, muscle belly lengthening velocity and acceleration [35], and 1 focused on the relationship between optimal fascicle isokinetic dynamometer and fascicle length during gait [61].

## 4. Discussion

This scoping review aimed to provide a comprehensive overview of how MSK modeling and USI are used during 3DGA in individuals with neuromotor impairments, and to examine their clinical relevance. Overall, MSK modeling was more commonly used than USI (MSK: n=45 studies, USI: n=8 studies) and used to assess clinical outcome (e.g., effects of surgical procedures or botulinum toxin injections), whereas USI was predominantly used to provide biomechanical insights into muscle and tendon behavior during gait (e.g., fascicle length, AT length).

### 4.1. Current use and clinical contribution

#### 4.1.1. Comparisons with controls

Results of this scoping review suggested that MSK modeling and USI have enabled the assessment of MTU length and lengthening velocity across gait phases, and the identification of abnormal MTU kinematics in pathological gait. For instance, using MSK modeling, Jonkers et al.(2006) observed a premature maximal rectus femoris MTU lengthening velocity occurring during the stance phase in children with CP with increased spasticity, whereas in CON, this peak occurred just before toe-off, highlighting altered muscle timing as a potential contributor to stiff knee gait [39]. Using USI, Barber et al.(2017) highlighted that, in children with CP, plantar flexor fascicles frequently lengthen until reaching their maximum during eccentric contractions, suggesting a compensatory reliance on the passive mechanical properties of the MTU, possibly contributing to increased metabolic cost during gait due to a sub-optimal use of the elastic properties of the tendon [34]. The impaired muscle-tendon interactions may be related to deficits in strength and selective motor control [34], but alterations in passive muscle and tendon properties were also proposed as contributing factors. All studies using USI observed that plantar flexor fascicles of children with CP consistently exhibit eccentric behavior during the stance phase, despite variations linked to gait patterns (e.g. crouch or equinus gait), [4,34,35,61–63], in contrast to CON whose fascicles primarily contract isometrically [84]. Kalsi et al.(2016) raised concerns that such eccentric muscle behavior, particularly known for its potential to induce muscle damage [85], might contribute to muscle mass loss in the plantar flexors [63]. They suggests that repeated muscle injury, combined with a potential reduction in the number of muscle satellite cells [86], could progressively reduce both muscle volume and force-generating capacity [63]. Given that eccentric damage typically occurs at high muscle lengths and contraction velocity [87,88], future studies should focus on determining whether these conditions are met across different pathological gait patterns.

#### 4.1.2. The effect of surgical or pharmacological interventions

Only MSK modeling studies have investigated the effect of surgical or pharmacological interventions on MTU length and/or kinematics. Wren et al.(2004) observed that gastrocnemius recession effectively restored both static and dynamic MTU length in children with equinus gait, particularly correcting preoperatively short muscle lengths [50]. Other studies have used MSK modeling to assess the outcomes of hamstring lengthening with more mixed results. For instance, Salami et al.(2019) reported a significant short-term increase in hamstrings MTU length following lengthening surgery in children with CP but noted that this effect was only maintained in children whose hamstrings were preoperatively short but not slow [30]. In contrast, Park et al.(2009) reported that while distal hamstring lengthening improved knee kinematics, it did not result in significant changes in MTU length during gait, suggesting MSK modeling may not reliably reflect functional improvements or surgical needs [52]. The discrepancy between findings may be partly explained by the fact that the latter did not account for preoperative muscle characteristics, such as muscle shortness or slowness, which were shown to be critical in predicting post-surgical changes in MTU length [30,41,50].

Following botulinum toxin type A injections, Kim et al.(2020) quantified changes in semimembranosus MTU kinematics in children with CP with flexed knee gait, revealing a significant increase in MTU lengthening velocity during swing phase without changes in absolute MTU length, suggesting a primarily dynamic rather than structural effect of the intervention [31]. Also, Thompson et al.(1998) observed that botulinum toxin injections produced repeatable increases in hamstring MTU length and excursion, particularly in muscles that were initially short, and contributed to improved knee extension during stance, highlighting the importance of identifying true muscle shortening prior to intervention [45].

#### 4.1.3. The effect of physical interventions

MSK modeling has been used to assess muscle excursion following gait training. DeVol et al.(2025) quantified changes in MTU lengths and lengthening velocity in children with CP following spinal stimulation combined with short-burst treadmill training, revealing that although joint kinematics improved, increases in MTU excursion were modest and only weakly correlated with reductions in spasticity [56]. However, no studies have examined how physical interventions, such as gait training, strength exercises or neuromuscular stimulation, affect MTU behavior and functional outcomes.

To date, no longitudinal study using USI has been conducted to assess muscle of MTU changes following physical interventions, highlighting a critical gap in the literature.

#### 4.1.4. Predictors of surgical outcomes

MSK modeling and USI have been used to explore predictors of surgical outcomes. These predictors are measurable parameters, such as MTU length, lengthening velocity, or timing of peak muscle stretch, that may predict the success or failure of a surgical intervention such as tendon lengthening. For instance, using MSK modeling, Arnold et al.(2006) have observed that hamstrings MTU length and kinematics can distinguish individuals who have “short” or “spastic” hamstrings, and help to determine if hamstrings lengthening can improve knee extension during walking [25]. Also, Desailly et al.(2011) have observed, in children with stiff knee gait, that a premature peak in rectus femoris length prior to surgery was associated with greater post-operative improvements, highlighting the potential of MTU kinematics as prognostic indicators.

Using USI, Harkness-Armstrong et al.(2021) reported in ITW individuals an adaptive shift in the muscle’s optimal length, leading to a more plantarflexed position on the force–length curve, which could partly explain the variable outcomes observed after MTU lengthening surgeries [61].

### 4.2. Methodological considerations and limitations

MSK modeling and USI allow detailed estimations of muscle and/or MTU geometry and/or kinematics but are limited by their labor-intensive nature and technical challenges in data acquisition and processing.

Regarding MSK modeling, one study has highlighted the importance of a consistent workflow [33]. The authors suggested using joint kinematics and MSK models that share matching segment reference frames and joint degrees of freedom, and to compare MTU lengths across groups only when the same modeling operations is applied. However, other sources of variability within the modeling pipeline remain largely unexplored. The effects of anatomical assumptions, such as predefined muscle paths or via-points, as well as scaling methods, including how marker weights influence model personalization [89], are still not well understood. These factors may significantly affect MTU length estimations, especially in populations with bone torsion such as individuals with CP and warrant further investigation to improve the robustness and reproducibility of MSK modeling outputs.

USI remains time-consuming, technically complex, and dependent on the assessor’s expertise [90], which can limit its clinical applicability and reproducibility. The manual or semi-automated processing of USI data, such as fascicle or tendon tracking, requires significant training and time investment, introducing variability between assessors [91]. To overcome these limitations and promote broader clinical integration, recent efforts have explored automation through deep learning-based approaches. For instance, python packages (e.g., DL_Track_US)[92] and neural network algorithms [93] have demonstrated promising results in automating the analysis of MTJ displacements and fascicle tracking during dynamic tasks. These advances may significantly reduce processing time, contribute to a standardized practice and therefore facilitate the clinical adoption of USI in 3DGA.

A study included in this scoping review has evaluated whether wearing an ultrasound device affected gait pattern, generally reporting non-significant gait alterations, which tend to be lower when the probe is placed on the mid-muscle fascicles [65], thereby supporting the feasibility of in-motion data collection. However, this study included children with a high functional level (i.e. GMFCS I-II). Patients with more severe disabilities (e.g. excessive hip adduction or internal rotation) may be more functionally constraint by an ultrasound probe. Finally, structural changes in muscles and connective tissues following a neurological lesion, such as increased deposition of connective tissue and intramuscular fat, and reduced skeletal muscle mass [94], are likely to affect ultrasound image quality. These alterations typically result in increased tissue echogenicity, which may impair the visualization of muscle architecture (e.g., fascicles, aponeuroses) and compromise measurement accuracy [95]. However, none of the included studies reported whether participants or ultrasound images were excluded due to poor image quality or excessive movement limitations. It is possible that this reflects either lack of occurrence or simply under-reporting. The fact that such methodological details are omitted makes it difficult to assess the potential for selection bias, which can affect both internal validity and reproducibility [96].

### 4.3. Recommendation for future studies

As the field of MSK modeling and USI continues to evolve, several directions emerge from this scoping review to enhance the applicability and inclusiveness of MTU assessment methods in both clinical and research contexts.

First, most studies focused on individuals with CP (n=36/50 studies), likely because CP is one of the most common neuromotor disorders affecting gait from childhood, and MTU abnormalities are a key feature of its clinical presentation. The gait patterns complexity and variability, combined with the frequent need for surgical or therapeutic interventions, make this population particularly relevant for in-depth monitoring of the MTU behaviour during gait. However, other neuromotor conditions such as stroke, multiple sclerosis, sarcopenia, or muscular dystrophy remain underexplored. These conditions may not yield comparable results, as they occur in fully developed musculoskeletal systems, whereas CP is characterized by early-onset neurological lesions combined with growth-related adaptations [97,98]. Thus, future studies should aim to include a broader range of pathologies. For instance, research involving stroke survivors could offer insights into spasticity and asymmetrical motor recovery; studies on individuals with multiple sclerosis may help clarify the interaction between spasticity, fatigue, and gait instability; and investigations in muscular dystrophies could support the monitoring of progressive muscle degeneration and the development of strategies to preserve mobility.

Second, studies should provide more detailed reporting of their experimental protocols to enhance reproducibility and facilitate inter-study comparisons. For instance, half of studies (n=25/50 studies) omitted reporting participants’ footwear during 3DGA [4,27,30,33,36,38,39,41–45,47–51,55,57,60,66,67,69–71], despite evidence indicating that footwear can significantly alter gait kinematics [99] and, consequently, MTU lengths. Similarly, some studies referred to the vastii muscles without specifying which one (vastus lateralis, medialis, or intermedius) was analyzed [40,67]. Moreover, the walking speed, a critical parameter influencing muscle and MTU kinematics, was often unreported (n=23/50 studies)[24,27,28,30,32,33,37,38,42–46,50–52,54,55,57,60,66,70,71].

Third, none of the included studies using USI were designed with a direct clinical objective or employed a longitudinal approach, limiting the clinical implications of USI at this stage. Future research using USI should investigate the effects of interventions, including strength training, botulinum toxin injections, or surgical procedures on the muscle and/or MTU during gait.

Fourth, only one study has investigated muscle-tendon interaction [34], despite evidences that muscle and tendon components can function semi-dependently during various functional tasks [100,101]. During walking, Achilles tendon passively store and release elastic energy, while muscles maintain nearly isometric contraction patterns [102]. Ultrasound-tracking of the MTJ has revealed this decoupling in vivo and highlighted its role in efficient force transmission and energy storage [102]. Future studies should investigate muscle–tendon interactions using in vivo tools like ultrasound, especially in pathological gait, to enable clinicians to design targeted interventions, such as high□strain eccentric training to modify tendon stiffness [103], or to optimize surgical or orthotic approaches that preserve beneficial tendon recoil.

### 4.4. Limitations

This scoping review presents several limitations. First, only studies published in English were considered to ensure full comprehension by all members of the review team. This may have introduced a language bias and limited the international scope of the evidence. Second, since the nature and heterogeneity of the included studies did not allow for a systematic review or meta-analysis, the analysis was descriptive and did not assess the strength or level of evidence. Finally, although the inclusion criteria were intentionally limited to MSK modeling and USI, currently the most widely used and emerging non-invasive methods for assessing muscle and/or MTU characteristics, other promising approaches such as elastography or real-time MRI were excluded.

## 5. Conclusion

Integrating MSK modeling and USI with 3DGA holds potential to refine diagnosis, guide targeted interventions, and improve functional outcomes in individuals with neuromotor impairments. This scoping review highlights that MSK modeling is more commonly used than USI during 3DGA and has been used to assess muscle and MTU characteristics, therapeutic outcomes, and predictors of surgical outcomes. Studies using USI are mostly cross-sectional, with limited clinical application. The predominant focus on CP individuals over other neuromotor disorders, along with inconsistent methodological reporting, a lack of longitudinal designs, and limited investigation into muscle–tendon interactions, hinders the current clinical applicability of these techniques.

## Supporting information

Supplementary file: Database Search Syntax

## Data Availability

All data produced in the present work are contained in the manuscript

