## Supplementary file: Database Search Syntax for "Experimental investigation of muscle-tendon unit geometry and kinematics in lower-limb muscles during gait: Current Applications and Future Directions – A Scoping Review"

**Database Search Syntax (Supplementary File)**

**PubMed**

*Domain concept:* Muskuloskeletal Modelling

1. "muscle modeling"
2. "musculoskeletal model*"
3. "human model*"
4. opensim
5. "biomechanical model*"
6. "muscle mechanics"
7. "SIMM"
8. "AnyBody"
9. "musculoskeletal simulation"
10. "computational modeling"
11. length*
12. 1-11 OR

*Domain concept:* Ultrasonography

13. ultrasound*
14. echograph*
15. sonograph*
16. ultrasonograph*
17. ultrasonic
18. "in vivo"
19. MRI
20. "magnetic resonance imaging"
21. 13-20 OR

*Domain concept:* Lower extremity

22. "lower extremit*"
23. "lower limb"
24. "leg"
25. "tibialis anterior"
26. "gastrocnemi*"
27. "soleus"
28. "triceps surae"
29. "vastus lateralis"
30. "vastus medialis"
31. quadriceps
32. "rectus femoris"
33. Semitendinosus
34. Semimembranosus
35. hamstring*
36. "biceps femoris"
37. gracilis
38. sartorius
39. psoas
40. "tensor fasciae latae"
41. "gluteus minimus"
42. "gluteus medius"
43. "gluteus maximus"
44. knee
45. hip
46. ankle
47. 22-46 OR

*Domain concept:* Gait

48. gait
49. walk*
50. treadmill
51. ambulat*
52. locomotion
53. 48-52 OR

*Domain concept:* Muscle Geometry

54. geometr*
55. size
56. volume
57. elongation
58. strain
59. architecture
60. length
61. "muscle-tendon length"
62. "muscle-tendon velocity"
63. "muscle-tendon unit"
64. "muscle-tendon interaction*"
65. "muscle-tendon junction"
66. "pennation angle"
67. "pinnation angle"
68. fascicle
69. interplay
70. behavior
71. behavior
72. 54-71 OR

*Domain concept:* Pathology

73. stroke
74. “cerebral palsy”
75. “spinal cord injur*”
76. hemiparesis
77. 73-76 OR

*Results:*

78. (12 OR 21) AND 47 AND 53 AND 72 AND 77 AND muscle*
79. limit 78 to [human subjects] NOT “systematic review” NOT “scoping review”

**Scopus**

*Results:*

80. (12 OR 21) AND 47 AND 53 AND 72 AND 77 AND muscle*
81. limit 80 to ([English language] AND [human subjects]) AND [Source: journal OR article]) NOT “systematic review” NOT “scoping review”

**ClinicalTrials**

*Condition/disease:*

82. stroke
83. “cerebral palsy”
84. “spinal cord injur*”
85. hemiparesis
86. 82-85 OR

*Accepts healthy volunteers:*

87. Yes

*Study type:*

88. interventional
89. observational
90. 88-89 OR

*Title:*

91. 1-11 OR
92. 13-20 OR

*Results:*

93. 86 AND 87 AND 90 AND (91 OR 92)
